# Immune profiling of *Mycobacterium tuberculosis*-specific T cells in recent and remote infection

**DOI:** 10.1101/2020.11.13.20230946

**Authors:** Cheleka A.M. Mpande, Virginie Rozot, Boitumelo Mosito, Munyaradzi Musvosvi, One B Dintwe, Nicole Bilek, Mark Hatherill, Thomas J. Scriba, Elisa Nemes, the ACS Study Team

**Affiliations:** South African Tuberculosis Vaccine Initiative, Institute of Infectious Disease and Molecular Medicine, Division of Immunology, Department of Pathology, University of Cape Town, South Africa

## Abstract

**Background:** Recent *Mycobacterium tuberculosis* (M.tb) infection is associated with a higher risk of progression to tuberculosis disease, compared to persistent infection after remote exposure. However, current immunodiagnostic tools fail to distinguish between recent and remote infection. We aimed to characterise the immunobiology associated with acquisition of M.tb infection and identify a biomarker that can distinguish recent from remote infection.

**Methods:** Healthy South African adolescents were serially tested with QuantiFERON-TB Gold to define recent (QuantiFERON-TB conversion <6 months) and persistent (QuantiFERON-TB+ for >1.5 year) infection. We characterized M.tb-specific CD4 T cell functional (IFN-γ, TNF, IL-2, CD107, CD154), memory (CD45RA, CCR7, CD27, KLRG-1) and activation (HLA-DR) profiles by flow cytometry after CFP-10/ESAT-6 peptide pool or M.tb lysate stimulation. We then assessed the diagnostic performance of immune profiles that were differentially expressed between individuals with recent or persistent QuantiFERON-TB+.

**Findings:** CFP-10/ESAT-6-specific CD4 T cell activation but not functional or memory phenotypes distinguished between individuals with recent and persistent QuantiFERON-TB+. In response to M.tb lysate, recent QuantiFERON-TB+ individuals had lower proportions of highly differentiated IFN-γ+TNF+ CD4 T cells expressing a KLRG-1+ effector phenotype and higher proportions of early differentiated IFN-γ-TNF+IL-2+ and activated CD4 T cells compared to persistent QuantiFERON-TB+ individuals. Among all differentially expressed T cell features CFP-10/ESAT-6-specific CD4 T cell activation was the best performing diagnostic biomarker of recent infection.

**Interpretation:** Recent M.tb infection is associated with highly activated and moderately differentiated functional M.tb-specific T cell subsets, that can be used as biomarkers to distinguish between recent and remote infection.

## Introduction

Tuberculosis remains a major global health problem, causing approximately 1.4 million deaths annually [1]. Despite being responsible for the most deaths due to a single infectious agent, only a minority (5-10%) of immunocompetent individuals infected with M.tb actually progress to disease. The risk of disease progression is not uniform in immunocompetent M.tb infected individuals, with the highest risk occurring within the first 2 years of primary (recent) M.tb infection [2–6]. By comparison, risk of tuberculosis in established, remote infection that has persisted for > 2 years is much lower [7]. Re-exposure of individuals with remote M.tb infection has been associated with a significantly lower rate of disease progression compared with exposure of individuals without prior infection [7,8]. Similarly, a non-human primate tuberculosis model has demonstrated that primary M.tb infection induces immunity that prevents re-infection or restricts bacterial growth upon re-infection [9].

Experimental M.tb infection in animal models has demonstrated that an early peak in M.tb burden occurs during acute infection. This is followed by immune-mediated reduction of bacterial replication that can persist during chronic infection for very long periods [10,11]. We postulated that recent M.tb infection in humans follows the same M.tb replication kinetic, albeit over a longer temporal period, resulting in a persisting “lifelong” low-grade infection in many individuals that facilitates maintenance of cellular immunity or, alternatively, eventual clearance of infection despite persistent immunological sensitisation [12]. In contrast, failure to control bacterial replication results in high and persistent mycobacterial burdens which are associated with disease progression. Since the magnitude and duration of *in vivo* T cell exposure to mycobacterial antigens are known to affect differentiation and functional capacity of CD4 T cells [13,14], we propose that different states of M.tb infection or disease will be associated with distinct T cell functional and differentiation states. Current tests for M.tb infection status such as tuberculin skin tests (TST) or IFN-γ release assays (IGRAs) measure T cell memory responses to M.tb antigens. A major limitation of these tests is that they do not reveal the duration of infection and, as a consequence, most immunology studies of M.tb infection have focused on remote M.tb infection. Our current knowledge of T cell immunity in the context of M.tb infection and disease is thus predominantly based on cross-sectional studies of individuals with remote M.tb infection or recent tuberculosis diagnosis, and very little is known about the T cell features associated with recent M.tb infection.

Upon antigen exposure, T cells become activated, differentiate and acquire functional properties. Expression of HLA-DR, a member of the MHC class II family typically expressed by antigen-presenting cells, is often used as a marker of T cell activation [15,16]. In the context of tuberculosis, studies have shown that disease, regardless of HIV status, is associated with significantly higher levels of T cell activation than M.tb infection, and that activation is reduced upon successful antimicrobial treatment [17–22]. These studies suggest that antigen-specific T cell activation could be used as a biomarker of microbial burden, and potentially infer *in vivo* antigen exposure during different stages of infection and disease. In addition, upon exposure to high antigen load CD4 T cells predominantly express IFN-γ, which is mostly associated with an effector memory T (T_E_) cell phenotype. Conversely, expression of TNF and IL-2 in the absence of IFN-γ is associated with stem cell memory (T_SCM_) and central memory (T_CM_) T cell subsets and exposure to low antigen load or antigen clearance [13,23]. Further, chronic duration of antigen exposure is associated with upregulation of the T cell senescence marker, KLRG-1 and T cell differentiation to a predominantly IFN-γ-monofunctional capacity [13,14]. These highly differentiated antigen-specific IFN-γ-monofunctional T cells have a lower capacity to contribute to M.tb clearance than polyfunctional Th1 cells, and are positively correlated with an increase in bacterial burden [24,25]. Based on these cytokine co-expression profiles we defined a single measure, which we termed *functional differentiation score* (FDS), that aims to infer the degree of T cell differentiation [14]. A higher M.tb-specific T cell FDS was associated with increased magnitude and duration of antigen exposure and T_E_ memory phenotypes [14].

TST and IGRAs have a low predictive value for tuberculosis [26]. Other functional assays that measure M.tb-specific T cell properties such as Th1 cytokine co-expression profiles [27,28], T cell differentiation (CD27, [29,30]) and T cell activation [17–22] have been proposed as new potential immunodiagnostic concepts that can distinguish between M.tb infection and disease. In line with this principle, potential biomarkers of recent M.tb infection based on proportions of TNF-only (IFN-γ-IL-2-) T_E_ (CD45RA-CCR7-CD127-) CD4 T cells or T cell proliferation, as well as a blood transcriptomic signature, have been described [31–33]. Halliday and colleagues demonstrated that the proportion of PPD-reactive TNF-only T_E_ CD4 T cells was highest in tuberculosis patients and lowest in individuals with remote M.tb infection, and distinguished between recent and remote M.tb infection with a sensitivity and specificity of 89% and 65%, respectively [31]. These studies all present evidence for potential biomarkers of tuberculosis risk. However, in-depth characterisation of immunological features associated with recent acquisition of infection that could contribute to an improved understanding of M.tb-specific immunobiology were not presented. The purpose of our study was to characterise the kinetics of M.tb-specific T cell features associated with recent and remote M.tb infection. We *postulated that differential in vivo antigen exposure during recent and remote M*.*tb infection is associated with distinct M*.*tb-specific T cell memory, functional and activation profiles*. More specifically, *we hypothesised that recent infection is associated with lower T cell differentiation and higher T cell activation compared to remote infection*. We thus aimed to identify *M*.*tb-specific T cell features that could distinguish between recent and remote M*.*tb infection*.

## Methods

### Study Design

We designed a retrospective study in adolescents to compare immune responses between recent and remote M.tb infection using cryopreserved peripheral blood mononuclear cells (PBMC).

#### Participants

South African adolescent participants were enrolled in observational studies approved by the University of Cape Town Human Research Ethics Committee (protocol references: 045/2005, 102/2017). Adolescents provided written, informed assent and their parents or legal guardians provided written, informed consent. IGRA testing with QuantiFERON-TB Gold In-Tube (QFT; Qiagen) was performed to measure M.tb infection and PBMC were collected at enrolment and at 6-monthly intervals during a 2-year follow-up in a subset of the cohort. See supplementary methods for detailed inclusion and exclusion criteria.

##### Definition of recent and remote M.tb infection

We defined two adolescent cohorts of recent M.tb infection based on serial QFT (and TST) results (**Supplementary Table 1**). Adolescents selected for the <u>biomarker discovery cohort</u> were defined as healthy participants with two negative QFT tests (IFN-γ < 0.35 IU/mL; assay performed and interpreted according to the manufacturer’s specifications) followed by two positive QFT tests (IFN-γ ≥0.35 IU/mL) 6 months apart over 1.5 years (**Figure 1A**). Recent QFT+ individuals selected for this cohort had at least one positive and negative QFT test result out of the QFT-uncertainty zone (0.2-0.7 IU/mL) defined by our research group [34]. Recently M.tb infected adolescents selected for the <u>tetramer cohort</u> were defined by a negative QFT (IFN-γ < 0.35 IU/mL) and TST negative (induration < 5mm) at baseline, followed by at least three positive QFT tests and one positive TST result (**Figure 1B**) [35]. All participant had quantitative QFT responses outside of the QFT uncertainty zone.

**Table 1:** AUCROC comparison of potential biomarkers of recent M.tb infection.

| CD4 T cell Subset 1 |  |  | CD4 T cell Subset 2 |  |  | Roc test<br>p-value |
| --- | --- | --- | --- | --- | --- | --- |
| Subset | Stim | AUCROC<br>(95% CI) | Subset | Stim | AUCROC (95% CI) |  |
| HLA-DR % of IFN-<br>γ+ | CFP-10/<br>ESAT-6 | 0.94<br>(0.83-1.00) | FDS | M.tb<br>lysate | 0.72 (0.56-0.88) | **0.018** |
|  |  |  | KLRG- 1+ IFN-γ+TNF+IL-2- T<sub>E</sub> |  | 0.74 (0.58-0.90) | **0.035** |
| HLA-DR % of<br>IFN-γ+TNF+ |  | 0.94<br>(0.84-1.00) | FDS |  | 0.72 (0.56-0.88) | **0.014** |
|  |  |  | KLRG- 1+ IFN-γ+TNF+IL-2- T<sub>E</sub> |  | 0.74 (0.58-0.90) | **0.030** |
| HLA-DR % of Th1<br>Cyt+ |  | 0.92<br>(0.81-1.00) | FDS |  | 0.72 (0.56-0.88) | **0.039** |
|  |  |  | KLRG- 1+ IFN-γ+TNF+IL-2- T<sub>E</sub> |  | 0.74 (0.58-0.90) | 0.062 |

**Figure 1:**
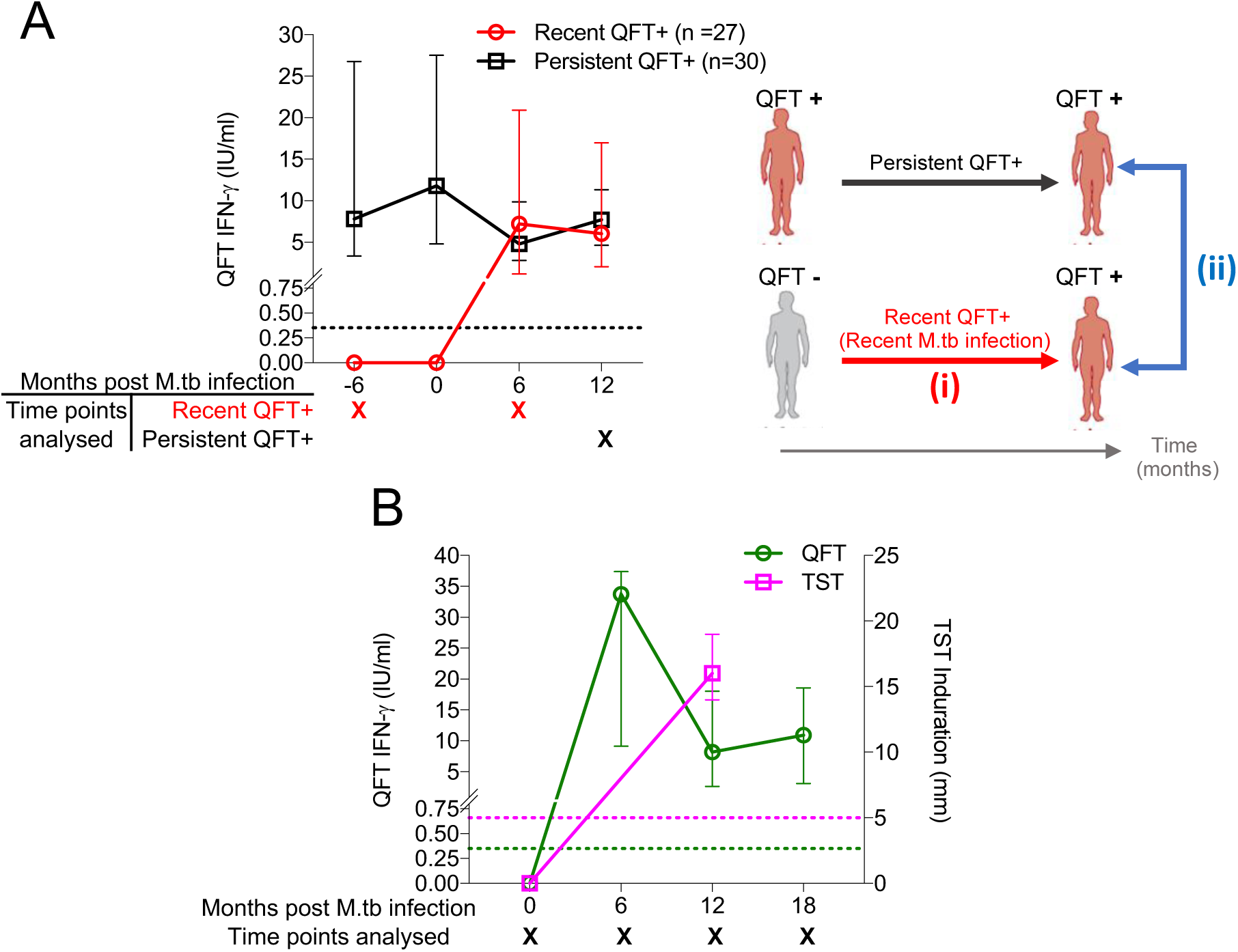
Longitudinal QFT (and TST) kinetics in the biomarker discovery and tetramer cohorts. (A) M.tb infection was defined using longitudinal (A) QFT (only) results for the biomarker discovery cohort [recent QFT+ (red symbols) and persistent QFT+ (black symbols)], and (B) both QFT (green) and TST (magenta) results for the tetramer cohort. Longitudinal QFT+ and TST results are depicted as median with an interquartile range (IQR). Dotted lines indicate the assay positivity cut-offs. Only samples collected at time points marked with “X” were analysed in this study. The image in (A) depicts comparisons that were performed on the biomarker discovery cohort: (i) QFT-vs recent QFT+ and (ii) recent vs persistent QFT+.

Remote (persistent) M.tb infection was defined in healthy participants by 4 consecutive QFT positive tests (two of which had to be > 0.7 IU/mL) 6 months apart over 1.5 years (<u>biomarker discovery cohort</u>, **Figure 1**). Here we included results from all visits from the tetramer cohort, while results presented for the biomarker discovery cohort were from the first QFT- and the first QFT+ visit (1 year apart) for recent QFT+ participants, and fourth QFT+ visit for persistent QFT+ participants (**Figure 1**).

### Protocols used to detect M.tb-specific CD4 T cells

M.tb-specific CD4 T cells were detected using either PBMC intracellular cytokine staining (ICS) or major histocompatibility complex (MHC) class II tetramer staining and flow cytometry. Detailed PBMC-ICS stimulation protocol is outlined in the *Supplementary Methods*. Briefly, cryopreserved PBMC were stimulated with peptide pools spanning full length CFP-10/ESAT-6, M.tb-specific antigens used in the QFT assay, or M.tb lysate (H37Rv) for 18 hours (biomarker discovery cohort). Cells were then stained with antibodies (**Supplementary Table 2**) and analysed by flow cytometry. For the ICS assay M.tb-specific cells were defined by cytokine expression (**Supplementary Figure 1 A**).

For the tetramer cohort, human leukocyte antigen (HLA)-typing was performed to determine which tetramers could be used to measure M.tb-specific T cells. Briefly, DNA was extracted from PBMC to determine high resolution HLA class I and II genotypes by polymerase chain reaction (PCR) using sequence-specific primers [36]. HLA allele ambiguities were resolved by allele-specific DNA sequencing [14,37]. For M.tb-specific MHC class tetramer staining the previously described protocol was followed [35]. Briefly, thawed cryopreserved PBMC were stained with anti-CCR7 for 30 minutes at 37°C, washed and then stained with 2μg/mL MHC class II tetramers (**Supplementary Table 3**) for 1 hour at room temperature. Cells were then washed and stained with surface marker antibodies (**Supplementary Table 4**) and analysed by flow cytometry (**Supplementary Figure 1 B**).

### Data Analysis

#### Biomarker discovery cohort analysis pipeline

We focused our analyses on CD4 T cells because frequencies of CD8 T cell responses were undetectable or not different to background in most individuals (data not shown). After conventional gating performed in FlowJo (**Supplementary Figure 1)**, we followed the analysis pipeline outlined in **Supplementary Figure 2** and described in the online methods. Briefly, expression of all possible combinations of functional responses (obtained by Boolean gates) were analysed using COMPASS [38], Pestle and SPICE [39]. In parallel, we utilised MIMOSA [40] to define responders (see below). We identified differentially expressed T cell subsets (including all possible combinations of functional and phenotypic markers) between two groups using CITRUS analysis (**Supplementary Figure 3-4)** in Cytobank on responders only [41]. Following CITRUS analysis, we exported CITRUS cluster FCS files, concatenated the files from all participants belonging to the same cluster and confirmed memory/cytokine cluster composition by manual re-gating in FlowJo.

**Figure 2:**
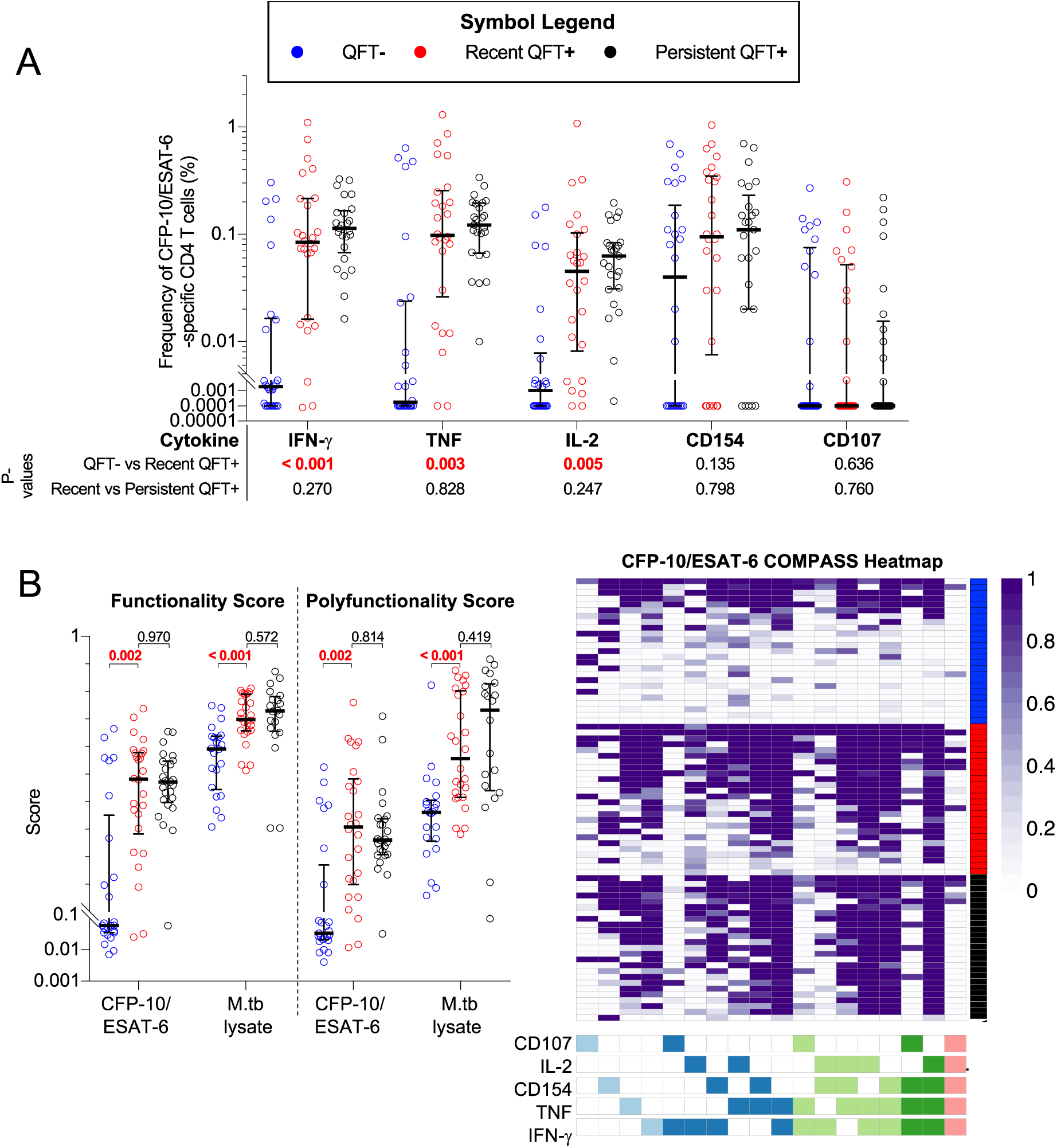
Recent QFT conversion induces of M.tb specific functional CD4 T cells with similar CFP-10/ESAT-6-specific CD4 T cell cytokine co-expression profiles as in persistent QFT+ individuals. (A) Frequencies of background subtracted CFP-10/ESAT-6-specific IFN-γ+ TNF+, IL-2+, CD154+ and CD107+ CD4 T cells and (B) COMPASS analysis [functionality scores (FS), polyfunctionality scores (PFS) and posterior probability heat map (CFP-10-ESAT-6 stimulation)] results of M.tb-specific CD4 T cells detected before (QFT**-**, blue, CFP-10/ESAT-6, n=26; M.tb lysate, n=22), after (recent QFT**+**, red, CFP-10/ESAT-6, n=26; M.tb lysate, n=26) and during persistent M.tb infection (persistent QFT**+**, black, CFP-10/ESAT-6, n=25; M.tb lysate, n=19). P-values were calculated using the Wilcoxon-signed rank for paired (QFT**-***versus* recent QFT**+**) or the Mann-Whitney U test for unpaired (recent *versus* persistent QFT+) comparisons and corrected for multiple comparison using the Benjamini–Hochberg method with a false discovery rate (FDR) of 0.05. P-values highlighted in **red, bold text** were considered significant.

**Figure 3:**
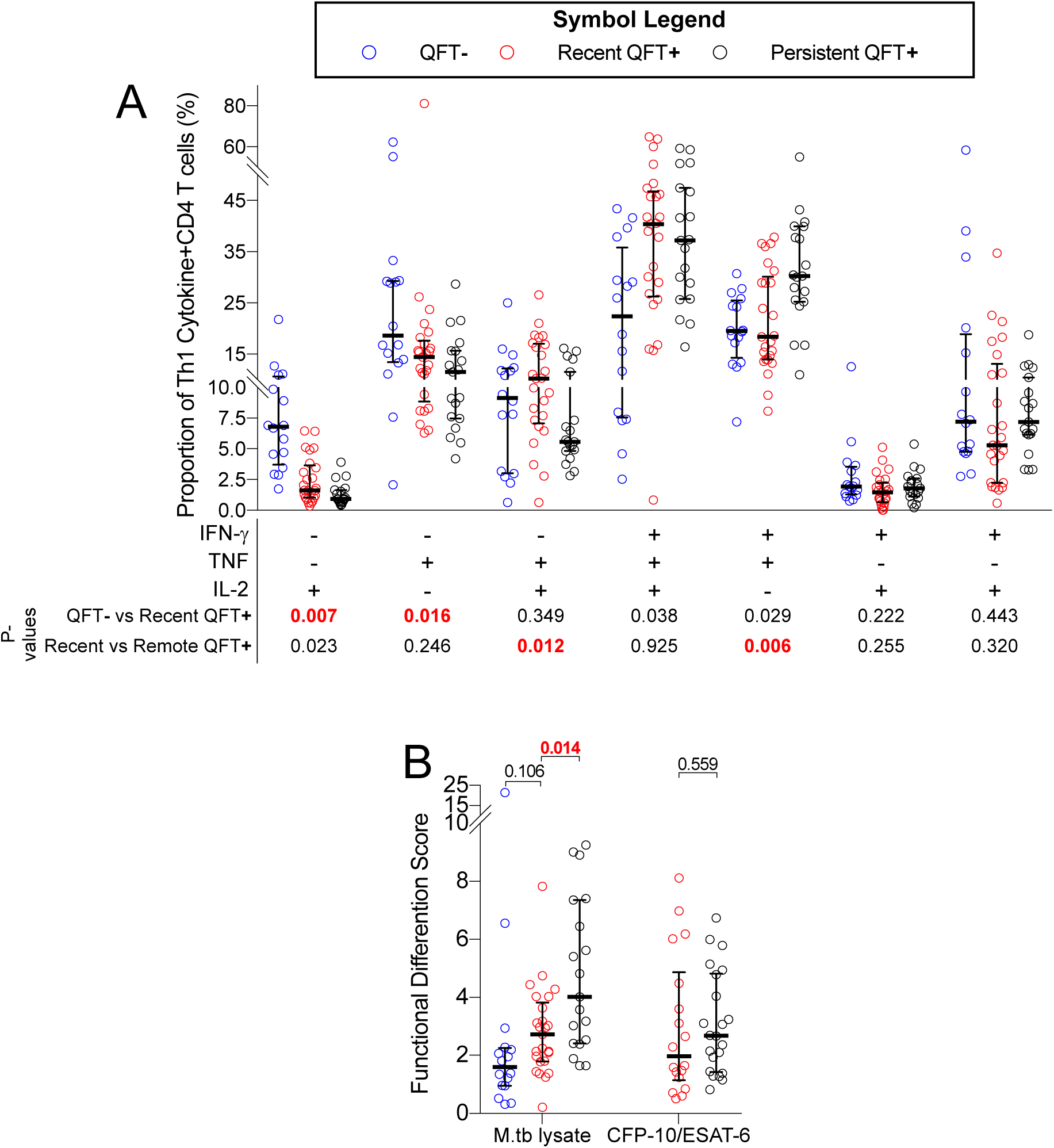
Recent QFT+ is associated with lower proportions of M.tb lysate-specific functionally differentiated CD4 T cell subsets than persistent QFT+. (A) Proportions of M.tb lysate-specific Th1 polyfunctional profiles detected before (QFT**-**, n= 16), after (recent QFT**+**, n=25) and during persistent M.tb infection (persistent QFT+, n=22) in responders only. (B) Functional differentiation scores of M.tb-specific CD4 T cells before (QFT**-**, M.tb lysate, n= 16), after (recent QFT**+**, CFP-10/ESAT-6, n= 18; M.tb lysate, n=25), and during persistent M.tb infection (black symbols, CFP-10/ESAT-6, n= 22; M.tb lysate, n=19) in responders only. P-values were calculated using the Wilcoxon-signed rank for paired (QFT**-***versus* recent QFT+) or the Mann-Whitney U test for unpaired (recent *versus* persistent QFT+) comparisons and corrected for multiple comparison using the Benjamini–Hochberg method with a false discovery rate (FDR) of 0.05. P-values highlighted in **red, bold text** were considered significant.

**Figure 4:**
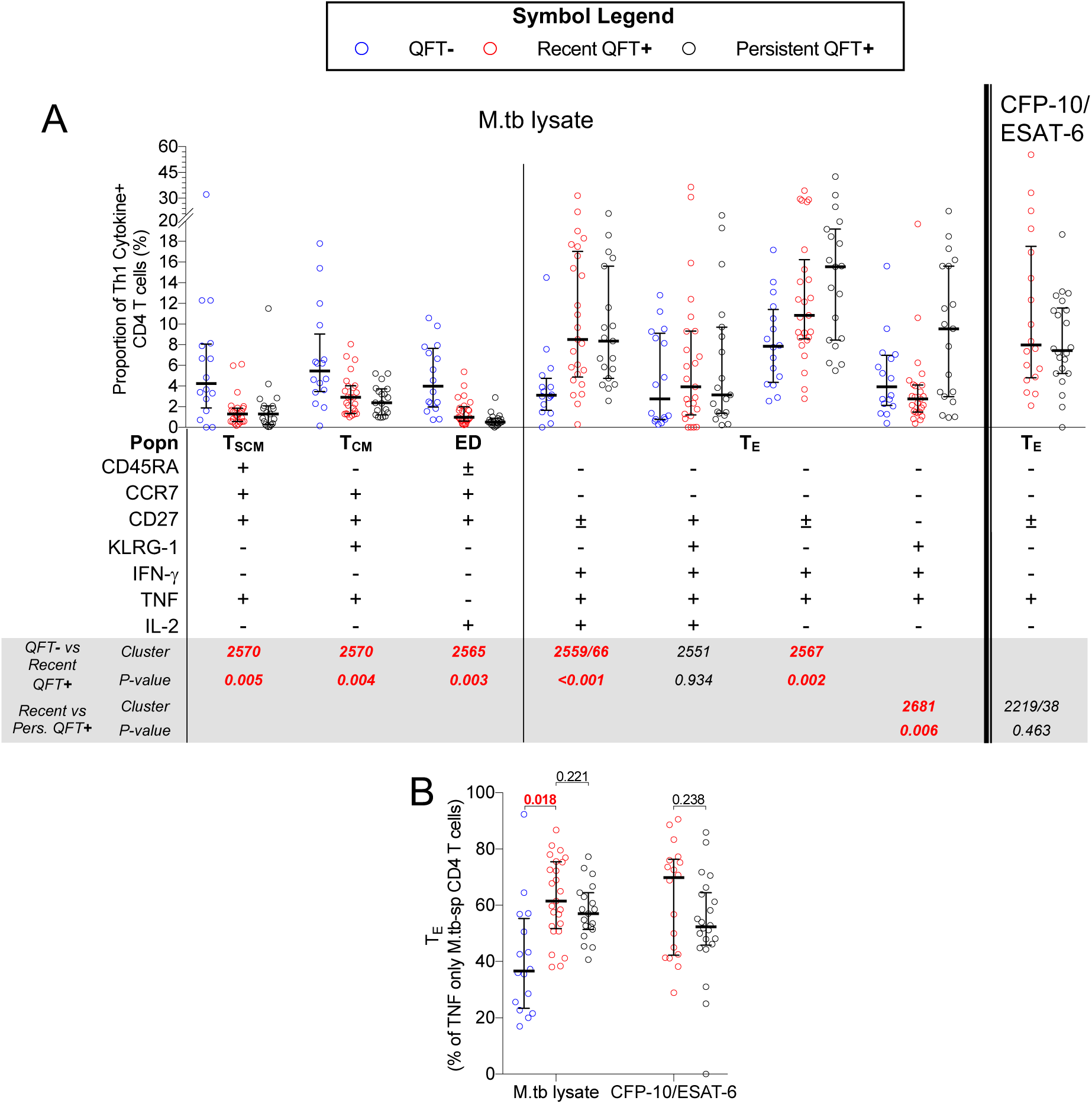
Induction of highly differentiated polyfunctional T_E_ cells upon recent QFT conversion at the expense of less differentiated IFN-γ-independent T_SCM_ and T_CM_ subsets. Proportions of (A) M.tb lysate-specific and CFP-10/ESAT6-specific memory/cytokine co-expression subsets before (QFT**-**, blue, M.tb lysate, n= 16), after (recent QFT+, red, CFP-10/ESAT-6, n= 18; M.tb lysate, n=25), and during persistent M.tb infection (persistent QFT+, black, CFP-10/ESAT-6, n= 22; M.tb lysate, n=19). The CITRUS cluster number corresponding to the manually gated populations is shown. (B) Proportions of T_E_ TNF only M.tb lysate-specific and CFP-10/ESAT-6-specific CD4 T cells detected before, after and during persistent M.tb infection (number of participants is as in A). Statistical analysis was conducted as in Figure 2, clusters and p-values highlighted in **red, bold text** were considered significantly different.

#### Biomarker discovery cohort responder definition

Responders were defined as individuals with frequencies of antigen-specific total Th1 cytokine+ (IFN-γ±TNF±IL-2±) CD4 T cells that were significantly higher [false discovery rate (FDR) ≤0.01, calculated by MIMOSA in R, version 1.21.0; https://github.com/RGLab/MIMOSA; [40] and at least 3-fold increase] than background. Non-responders were excluded from phenotypic marker expression analysis. We used this strict cut-off to ensure that cytokine responses in responders were truly antigen specific because analysis of activation, memory and/or functional phenotypes were performed using methods that would not account for non-specific signal using background subtraction.

#### Functional Differentiation Score

All possible cytokine co-expression T cell subsets were computed as posterior probabilities or summarised as a single functionality score (FS) or polyfunctionality score (PFS) for each individual using COMPASS [38]. We also calculated the FDS (Equation 1 [14]) for responders only.

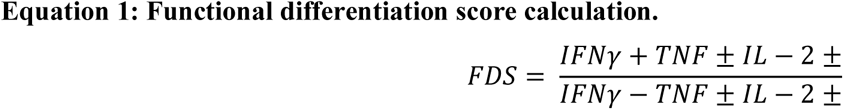

### Statistical analysis

Statistical analyses were performed using R, R Studio or GraphPad Prism v7. We applied Wilcoxon signed-rank and Mann-Whitney U statistical tests for paired and unpaired comparisons, respectively. Bonferroni or Benjamini-Hochberg (FDR <0.05) methods were used to correct for up to 4 comparisons or more than 4 comparisons, respectively. In addition to this, functional, memory and activation features identified as significantly different between recent and persistent QFT+ individuals were tested for their diagnostic potential using receiver operating characteristic (ROC) curve analysis performed using the pROC package in R [version 1.15.3; https://web.expasy.org/pROC/;[42]]

## Results

### TST dynamics in the biomarker discovery cohort

M.tb infection status of individuals in the biomarker discovery cohort, on which the ICS assay was performed, was based on QFT only. However, we also had access to TST results for most of these individuals and observed 100% concordance between QFT and TST results in recent QFT+ individuals, and an equally high concordance between QFT and TST results in persistent QFT+ individuals: proportion of agreement = 88.14%; Kappa = 0.763 (data not shown).

### Recent QFT+ is associated with lower T cell functional differentiation compared to persistent QFT+

We aimed to determine functional properties associated with acquisition of infection, and how they compare to persistent QFT+. Frequencies of CFP-10/ESAT-6-specific CD4 T cells expressing IFN-γ, TNF, IL-2 or CD107 were undetectable or very low before QFT conversion, as expected (**Figure 2A**). By contrast, CD154-expresisng CD4 T cells were detected in more than half of the individuals at this time point (**Figure 2A**). QFT conversion was associated with a significant increase in frequencies of M.tb-specific IFN-γ, TNF or IL-2-expressing CD4 T cell responses (**Figure 2A**). CD107 or CD154-expresisng CD4 T cells did not change significantly after QFT conversion.

We then utilised COMPASS analysis to determine FS, PFS and posterior probabilities of antigen-specific responses for all possible cytokine-co-expression subsets associated with acquisition of M.tb infection (**Figure 2B**). As observed above, FS, PFS and probabilities of antigen-specific responses of CFP-10/ESAT-6-specific CD4 T cells were very low prior to M.tb infection (QFT-) in most individuals. These outcomes increased significantly upon recent QFT conversion to levels that were similar to those in individuals with persistent QFT+ reactivity (**Figure 2**). Probabilities of antigen-specific responses in persistent QFT+ individuals were universally high for subsets that co-expressed two, three or four functions among IFN-γ, TNF, IL-2 and CD154, but not CD107. We next investigated M.tb lysate-specific CD4 T cell responses kinetics associated with acquisition of infection. M.tb lysate contains antigens that cross-react with bacille Calmette-Guérin (BCG) vaccine and other non-tuberculous mycobacteria *sp*., facilitating detection of mycobacterial responses in BCG-vaccinated M.tb uninfected individuals (QFT-) and comparisons of T cell features before and after M.tb infection. Prior to M.tb infection we detected robust levels of M.tb lysate-specific CD4 T cells with relatively high FS and PFS, which increased to even higher levels upon infection (recent QFT+) that were in a similar range to those detected in persistent QFT+ individuals (Figure 2 B).

T cells are known to change their functional capacity depending on antigen burden. We recently defined a single measurement of functional differentiation, FDS, that we propose can distinguish between infections states associated with different levels of M.tb burden [14]. We thus compared the distribution of cytokine co-expressing Th1 subsets and FDS in individuals in the biomarker discovery cohort. Since the majority of CFP-10/ESAT-6-specific CD4 T cells detected prior to M.tb infection were low and did not meet our responder criteria, we focused our longitudinal analyses of functional changes on M.tb lysate-specific CD4 T cells (**Supplementary Figure 5**). Relative proportions of M.tb lysate-specific CD4 T cells expressing only IL-2 or TNF were higher prior to than after M.tb infection, while proportions of IFN-γ-TNF+IL-2+ and IFN-γ+TNF±IL-2± subsets were not different before and after M.tb infection (**Figure 3 A**). Higher proportions of IFN-γ-TNF+IL-2+ but lower proportions of IFN-γ+TNF+IL-2-M.tb lysate-specific CD4 T cells were detected in recent QFT+ compared to persistent QFT+ individuals. This resulted in lower Th1 functional differentiation, measured by FDS, in recent QFT+ compared with persistent QFT+ individuals (**Figure 3 B**). FDS of CFP-10/ESAT-6-specific CD4 T cells between the two M.tb infection states were not different (**Figure 3 B**).

**Figure 5:**
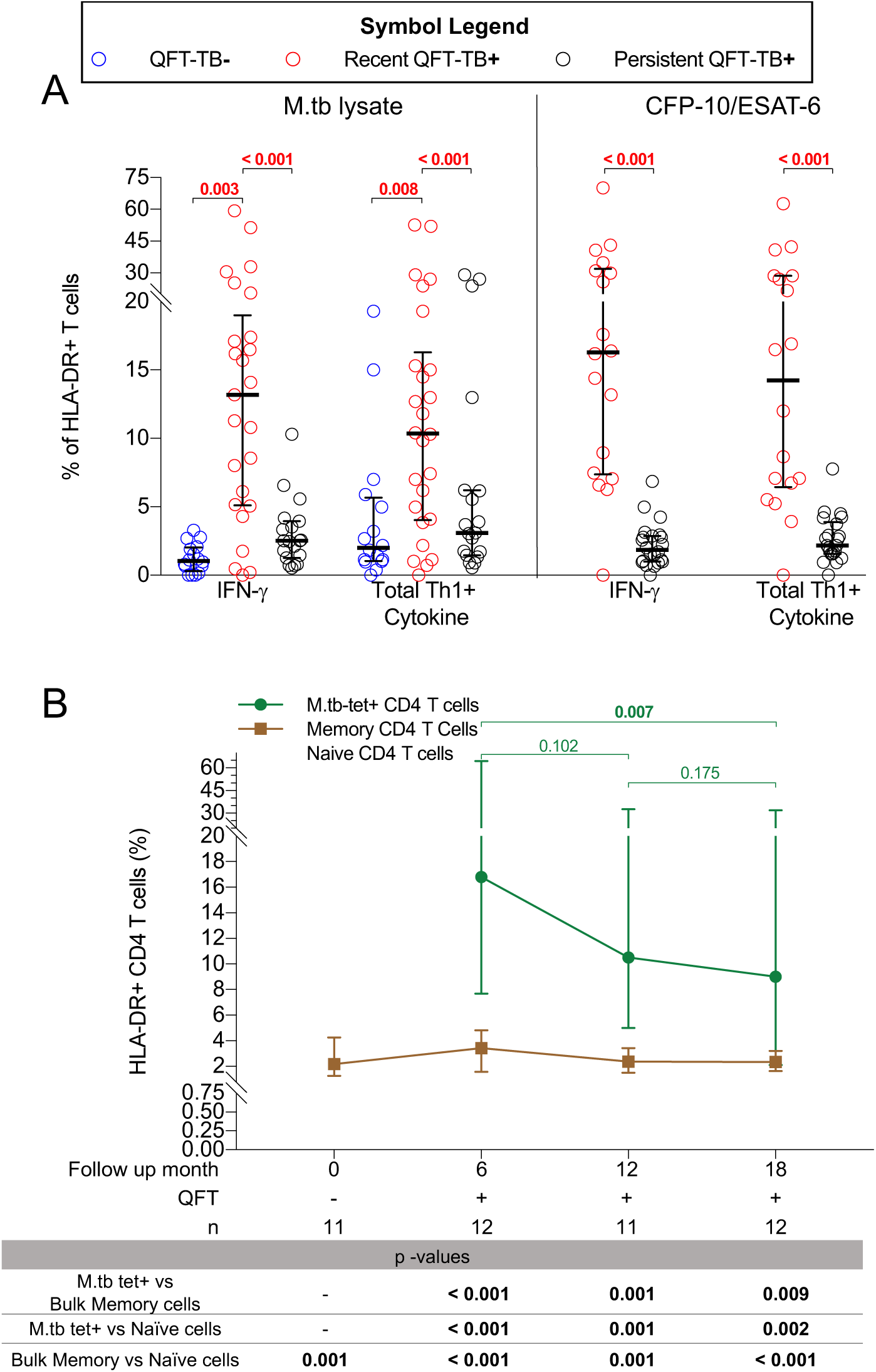
Recent QFT+ is associated with higher levels T cell activation than persistent QFT+. (A) Graphs depict proportions of HLA-DR+ CFP-10/ESAT-6- and M.tb lysate-specific IFN-γ+ or Total Th1 cytokine+ (IFN-γ±TNF±IL-2±) CD4 T cells before (QFT-, blue, M.tb lysate: n= 16), after (recent QFT+, red, CFP-10/ESAT-6: n=18; M.tb lysate: n= 25), and during persistent M.tb infection (persistent QFT+, black, CFP-10/ESAT-6: n=22; M.tb lysate: n=19). Statistical analysis was conducted as in Figure 2. (B) Proportions of HLA-DR+ M.tb-specific tetramer+ (green), bulk naïve (grey, CD45RA+CCR7+CD27+CD95-) and bulk memory (brown, non-naïve T cells) CD4+ T cells are represented by median (symbol) and IQRs (bars). P-values for (B) were calculated with the Wilcoxon signed-rank test. P-values <0.016 (for A) and < 0.05 (for B), highlighted in **bold** were considered significant.

### CFP-10/ESAT-6-specific memory T cell subsets induced by recent QFT conversion are maintained in persistent QFT+ individuals

We next aimed to determine if acquisition of M.tb infection is associated with a shift in M.tb-specific T cell memory phenotypes, as well as compare the distribution of M.tb-specific memory phenotypes between recent and persistent QFT+ individuals. Analysis of T cell memory markers expressed by M.tb lysate-specific CD4 T cells showed that recent QFT conversion was associated with a decrease in KLRG-1-T_SCM_ (CD45RA+CCR7+CD27+) and T_CM_ (CD45RA-CCR7+CD27+) cells, and an increase in KLRG-1-T_E_ (CD45RA-CCR7-CD27-) cells compared to the pre-infection (QFT-) time-point, indicative of increased antigen exposure (**Supplementary Figure 6 A**). Memory phenotypes of M.tb-specific responses, regardless of stimuli, were not different between recent and persistent QFT+ individuals, with transitional memory (T_TM_, CD45RA-CCR7-CD27+) and T_E_ cells being the predominant T cell memory subsets (**Supplementary Figure 6 B**).

**Figure 6:**
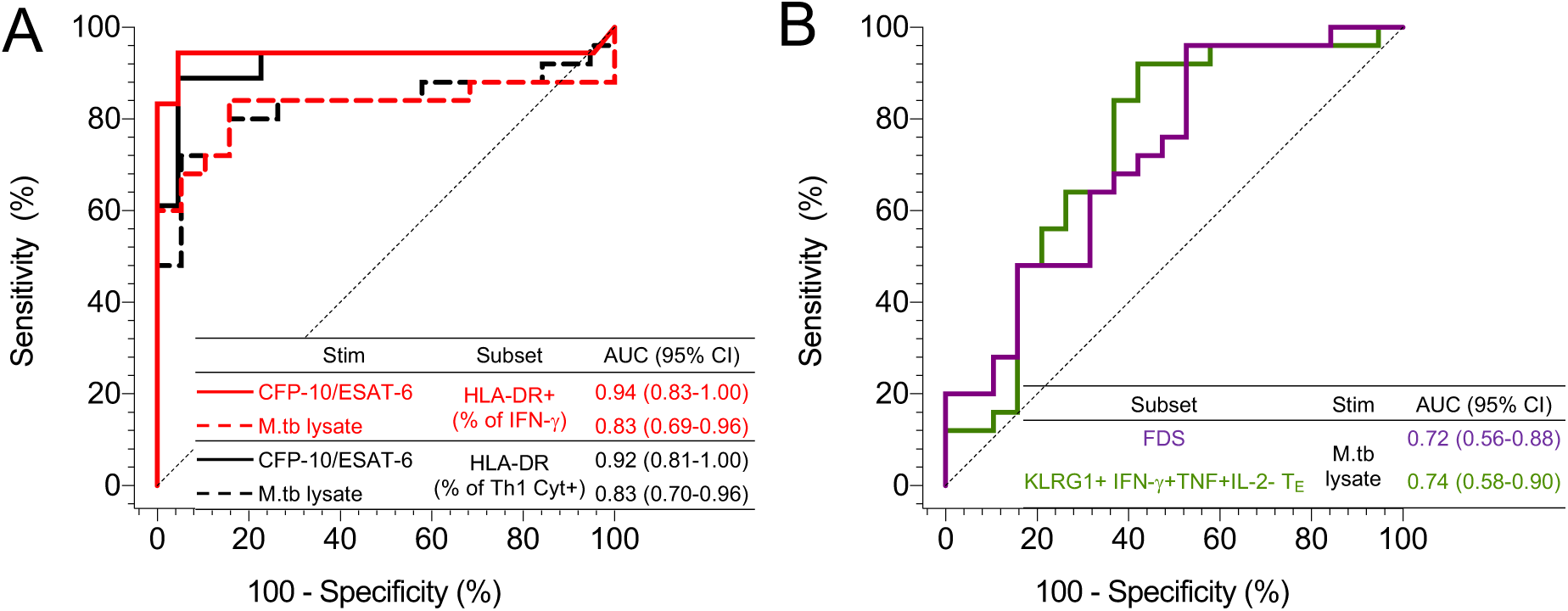
T cell activation is a biomarker of recent M.tb infection. ROC analysis of the diagnostic potential of M.tb-specific CD4 T cells subsets: (A) IFN-γ+ and Th1 cytokine+ T cell activation and (B) polyfunctional/memory subsets. All populations are significant defined based a 95% confidence intervals (CI) that do not cross 0.5.

Results above clearly illustrate a relationship between cytokine expression and memory phenotypes and suggest that combined analysis of these features may be informative. We therefore used CITRUS to identify cell subsets that were expressed at significantly different levels upon acquisition of M.tb infection, and between recent and persistent QFT+ individuals, based on expression of CD45RA, CCR7, CD27, KLRG1, IFN-γ, TNF and IL-2. We did not include CD107 and CD154 in this analysis, due to high non-specific, background expression of these markers. CXCR3 and HLA-DR were also excluded from this analysis because neither marker contributed to cell cluster definition during the optimisation of the CITRUS workflow. Composition of each differentially expressed T cell cluster, identified by CITRUS (***supplementary methods***), was confirmed by manual gating in all responders.

Compared to the pre-infection (QFT-) timepoint, recent QFT+ conversion was associated with a decrease in M.tb lysate-specific, early-differentiated IL-2-only T_SCM_/T_CM_ (cluster 2565) and TNF-only T_SCM_/T_CM_ (cluster 2570) cell subsets, and an increase in proportions of IFN-γ+TNF+IL-2+ (cluster 2559/66) and IFN-γ+TNF+ T_E_ (cluster 2567) cells (**Figure 4 A**). We also observed lower proportions of M.tb lysate-specific KLRG-1+IFN-γ+TNF+IL-2-(cluster 2681) T_E_ cells in recent QFT+ compared to persistent QFT+ individuals (**Figure 4 A**). Despite detecting differences in abundance levels of CITRUS clusters 2551 (QFT-*versus* recent QFT+) and 2219/43 (recent *versus* persistent QFT+), proportions of clusters were not different when manually gated (**Figure 4 A)**.

To the best of our knowledge, only one biomarker of recent M.tb infection, relative proportions of TNF-only (IFN-γ-IL-2-) T_E_ (CD45RA-CCR7-CD127-) CD4 T cells, has previously been reported as a diagnostic biomarker for differentiating between recent and remote M.tb infection [31]. We therefore aimed to evaluate the performance of this biomarker in our cohort. Because our flow cytometry panel did not include CD127, we defined this subset as TNF-only, CD45RA-CCR7-T_E_ memory cells. Acquisition of M.tb infection (QFT-*versus* recent QFT+) was associated with an increase in proportions of TNF-only T_E_ memory cells, however, unlike results presented by Halliday and colleagues [31], proportions of these cells, regardless of stimulus, were not different between recent and persistent QFT+ individuals (**Figure 4 B**).

These results confirm that recent QFT+ was associated with lower proportions of CD4+ IFN-γ-T_SCM_, T_CM_ and early differentiated memory cells, and higher proportions of activated and polyfunctional IFN-γ+ T_E_ cells, compared to the respective QFT-time point in each individual. Memory and functional profiles induced upon early infection were sustained at stable proportions once M.tb infection is established, with the exception of highly differentiated KLRG-1+IFN-γ+TNF+ T_E_ cells, which were more abundant during persistent QFT+ compared to recent QFT conversion.

### Higher levels of T cell activation in recent QFT+ than persistent QFT+ individuals

We next investigated T cell activation kinetics associated with acquisition of M.tb infection, and compared individuals with recent QFT conversion to those with persistent QFT+ status. Acquisition of M.tb infection (QFT-*versus* recent QFT+) was associated with a significant increase in both activated (HLA-DR+) IFN-γ+ and total Th1 cytokine+ M.tb-specific CD4 T cells (Figure 5A), which were also higher in recent QFT+ than in persistent QFT+ individuals (**Figure 5A**).

Since *in vitro* stimulation may affect expression of activation markers, we aimed to confirm T cell activation kinetics using MHC-class II tetramer staining, which detects antigen-specific cells directly *ex vivo* and does not require stimulation, on a different cohort of recently M.tb infected individuals [35]. Proportions of HLA-DR+ activated M.tb-tetramer+ CD4 T cells were highest at the first QFT+ time point (month 6 since last QFT negative test, when M.tb-tetramer+ cells were detectable) and markedly decreased by 1 year since QFT conversion (**Figure 5B**). Notably, the activation status of bulk memory and naïve CD4+ T cells were unchanged during the follow-up period. M.tb-tetramer+ CD4 T cells were also significantly more activated than bulk memory CD4 T cells, and bulk naïve CD4 T cells were significantly less activated than bulk memory CD4 T cells (**Figure 5 B**). These results demonstrate the robustness of T cell activation as a potential biomarker to distinguish between recent and persistent QFT+ individuals.

### T cell activation is a superior biomarker of recent M.tb infection than T cell differentiation

Finally, we investigated the diagnostic performance of functional, memory and activation profiles that were significantly different between recent and persistent QFT+ individuals in our dataset. We performed ROC analysis to assess the performance of HLA-DR+ IFN-γ+ and Th1 cytokine+ M.tb-specific CD4 T cells (regardless of stimulus), M.tb-lysate-specific FDS, and proportions of KLRG-1+ IFN-γ+TNF+IL-2-T_E_ (CD45RA-CCR7-CD27-) CD4 T cells to discriminate between recent and persistent QFT+ individuals. We did not perform ROC analysis on differentially expressed CD4 T cell cytokine co-expression profiles (IFN-γ-TNF+IL-2+, IFN-γ+TNF+IL-2-, IFN-γ+TNF-IL-2-) because these were already included in the definition of either the FDS or KLRG-1+ IFN-γ+TNF+IL-2-T_E_ cell population. Proportions of HLA-DR+ M.tb-specific (IFN-γ+ or Th1 cytokine+) CD4 T cells demonstrated very good discrimination between recent and persistent QFT+ individuals, achieving AUCs above 0.8 (**Figure 6 A**). By comparison, the highly differentiated KLRG-1+ IFN-γ+TNF+IL-2-T_E_ cell population and FDS demonstrated moderate discriminatory performance; AUCs for these comparisons were below 0.8 (**Figure 6 B**).

We also compared the discriminatory performance between recent and persistent QFT+ of CFP-10/ESAT-6-specific or M.tb lysate-specific HLA-DR+ IFN-γ+TNF+ CD4 T cells (Supplementary Table 7), an adaptation of a previously published biomarker with excellent discrimination between tuberculosis cases and healthy controls and which also showed promise as a tuberculosis treatment monitoring biomarker since it decreased significantly upon successful tuberculosis therapy [20]. Biomarkers based on HLA-DR expression by CFP-10/ESAT-6-specific CD4 T cells had significantly higher AUCs than those based on cytokine co-expression profiles or memory marker expression on M.tb lysate-specific CD4 T cells (**Table 1**). On the other hand, discriminatory AUCs of biomarkers based on HLA-DR expression regardless of M.tb stimulus or M.tb-specific CD4 T cell denominator were not different (p-value > 0.05), suggesting that neither subset had superior diagnostic potential over the other (Supplementary Table 7). Additionally, AUCs of biomarkers defined on M.tb lysate-specific activation and functional differentiation were all similar (p-value > 0.05). Thus, based on these results, M.tb antigen-specific T cell HLA-DR expression was identified as the best biomarker of recent M.tb infection in this dataset.

## Discussion

The risk of disease progression in M.tb infected individuals, identified by a positive IGRA or TST, is highest within the first 1-2 years after acquisition of M.tb infection [2,6]. Identification of individuals who recently became infected, and who would thus benefit most from tuberculosis preventive therapy, has only been possible with serial IGRA or TST testing. We performed in-depth analyses of M.tb-specific CD4 T cell function, activation and memory phenotypes to identify immunological differences between individuals who recently acquired infection and those with persistent (remote) infection. Three major points emerged from our study: 1) The acute response to M.tb-infection in humans was associated with highly activated M.tb-specific CD4 T cells that are moderately differentiated; 2) During remote infection M.tb-specific T cells were not activated but showed a phenotype consistent with long-term antigen exposure without evidence of exhaustion; 3) Among all the differences in M.tb-specific T cell phenotype and functions, HLA-DR expression had the highest diagnostic performance to distinguish between recent and remote infection.

We showed that recent acquisition of M.tb infection (QFT conversion from negative to positive) was associated with an increase in frequencies of Th1 (IFN-γ, IL-2 and TNF) cytokine-expressing CD4 T cells to levels in range with those detected in individuals with remote M.tb infection. We did not observe marked differences in cytokine co-expression profiles between recent and remote infection and therefore focused our analysis on combinations of Th1 cytokine co-expression (polyfunctional) profiles and T cell differentiation. As typical for T cell responses during acute and chronic infection [13], we observed a predominance of M.tb-specific highly activated cells and T_E_ cells upon recent acquisition of infection. The only distinguishing features between recent and remote infection were higher M.tb-specific T cell activation and lower proportions of M.tb lysate-specific KLRG-1+ IFN-γ+TNF+IL-2-T_E_ cells in recently compared to remotely infected individuals, while CFP-10/ESAT-6-specific T cell functional and memory profiles were very similar. In addition to being used as a proxy for T cell activation, HLA-DR expression by T cells has been suggested to facilitate antigen presentation to other T cells during times of high antigen burden [15,16], thus providing an additional source of cells beyond professional antigen presenting cells that can induce *de novo* T cell priming. Here we present human evidence, in line with findings observed in murine models [10], that suggests M.tb antigen load is higher during the acute phase of infection compared to more established phases of infection, when bacterial load is at least partially controlled.

Murine studies have also demonstrated that chronic M.tb infection is associated with differentiation of CD4 T cells to an exhausted T cell memory phenotype that expresses high levels of KLRG-1 and IFN-γ-monofunctional capacity, and poor ability to contribute to antigen clearance [24,25]. In contract with these observations from mice, here we show that despite long-term QFT-positivity, M.tb-specific T cells in remotely infected individuals did not have a dominance of KLRG-1+ IFN-γ-monofunctional exhausted T cells, nor did they show elevated activation consistent with persistently high antigen load. Instead, the activation levels of M.tb lysate-specific CD4 T cells during remote M.tb infection were not different to those observed before M.tb infection, suggesting little or no *in vivo* antigen exposure during remote M.tb infection. This may suggest that clearance of M.tb is rapid and usually occurs within 2 years in individuals who do not progress to disease.

Under this scenario, immune responses detected in most remotely infected individuals represent immunological memory, and not necessarily ongoing M.tb replication [2]. Alternatively, if remotely infected individuals truly harbour M.tb, as suggested by the higher functional differentiation of M.tb-specific T cells compared to recent infection, then levels of M.tb replication are likely low and stable, resulting in low T cell activation and a memory-functional phenotype not characteristic of T cell exhaustion. It is likely that persistent QFT+ status in humans represents a mixture of these scenarios.

Finally, we showed that of all the immunological features that were different between recent and remote M.tb infection, T cell activation was the best biomarker for differentiating between infection states. Overall, the T cell activation biomarker appears very robust, regardless of mycobacterial antigen specificity (M.tb lysate or CFP-10/ESAT-6) or the assay used for detection (PBMC-ICS, tetramer staining or, as previously reported, whole blood stimulation [17–22]). This study focused on understanding T cell immunobiology during acquisition of M.tb infection and the discovery of potential biomarkers to distinguish between recent and remote M.tb infection. Development of a simplified assay to identify such individuals, including validation in a test cohort and performance as a marker of disease progression, are beyond the scope of this paper and were reported separately (Mpande, under review, and [43]). This biomarker has the potential to identify individuals who are at high risk of progression and should receive tuberculosis preventive treatment, a key priority highlighted in the recent Global Tuberculosis Report [1], and differentiate them from those who are at low risk of progression and could be spared from treatment.

A limitation of this study was the small sample size and relatively short duration of follow-up of recently infected individuals, none of whom progressed to disease during the study. This prevented us from demonstrating how immunity in recently infected individuals who either progress to disease or develop persistent M.tb infection differ. Ideally, the M.tb-specific T cell activation biomarker should be prospectively validated on a cohort of individuals who acquire M.tb infection and then progress to disease. This would be an extraordinarily large study to conduct, since on a population level only a minority of individuals who acquire M.tb infection progress to disease [44]. Further, limited sample availability prevented us from testing other recently described biomarkers, such as those based on T cell proliferation [32] or gene expression [33]. In summary, we characterised changes in M.tb-specific T cell immunity in different phases of infection and present evidence of a peak in T cell activation, probably reflecting high M.tb burden, during recent infection followed by unexhausted T cell functional and memory phenotypes during remote, non-progressing infection, consistent with control or clearance of M.tb [2].

## Data Availability

Data will be available upon publishing in a peer-reviewed journal.

## Contributors

MH, TJS and EN designed the study and raised funding; CM, BM and NB generated the data; OBD and MM generated critical tetramer reagents; CM analysed the data; CM, VR, MH, TJS and EN interpreted the results; CM, MM, VR, TJS and EN wrote the manuscript. All authors revised and approved the manuscript and are accountable for the work.

## Acknowledgments

We are grateful to the study participants and their families; the Cape Winelands East district communities, the Department of Education and the Department of Health; the SATVI clinical and laboratory teams; Thomas Hawn, Morten Ruhwald and Jason Andrews for valuable input on the study design.

