## Supplementary Methods for "Immune profiling of *Mycobacterium tuberculosis*-specific T cells in recent and remote infection"

ACS Study Team: Hassan Mahomed, Willem A. Hanekom, Fazlin Kafaar, Leslie Workman, Humphrey Mulenga, Rodney Ehrlich, Mzwandile Erasmus, Deborah Abrahams, Anthony Hawkridge, E. Jane Hughes, Sizulu Moyo, Sebastian Gelderbloem, Michele Tameris, Hennie Geldenhuys, Gregory Hussey.

#### Methodology

##### *Participants inclusion and exclusion criteria*

Two groups of adolescents were selected (Supplementary Table 1) from a large epidemiological study conducted in the greater Worcester area, Western Cape, South Africa, from July 2005 through February 2009 [1]. Healthy 12-18 year-old participants were enrolled at high schools. Those pregnant or lactating, and those who reported acute or chronic medical conditions resulting in hospitalization within 6 months prior to enrolment were excluded. HIV testing was allowed only in participants diagnosed with tuberculosis during follow-up, and those who were HIV-positive were excluded from those included in groups analysed in this study.

##### *PBMC stimulation and staining protocols*

*Stimulation*: Cells were thawed, rested for 4 hours and stimulated in R10 media [RPMI 1640 Media (Gibco), 10% Fetal Bovine Serum (FBS, Gibco), 1% L-glutamine (Gibco) and 1% penicillin-streptomycin (Gibco)] containing anti-CD107a with either no antigen (unstimulated, negative control), CFP-10/ESAT-6 peptide pool (15 mer peptides overlapping by ten amino-acids, 1µg/mL, GenScript Biotechnology), M.tb lysate (H37Rv 10µg/mL, BEI Resources) or Staphylococcal enterotoxin B [SEB (positive control), 1µg/mL, Sigma Aldrich]. Cells were incubated for 3 hours at 37°C with 5% $CO_2$, after which brefeldin A (BFA, 5µg/mL, Sigma Aldrich) and monensin (2.5µg/mL, Sigma Aldrich) were added and incubated for a further 15 hours.

*Staining* (**Supplementary Table 2**): Cells were treated with 1mL of 2mM Ethylenediaminetetraacetic acid [EDTA, Sigma Aldrich, in phosphate buffer saline (PBS), Lonza], followed by centrifugation. Supernatant was discarded. Cells were then resuspended in 2mM EDTA and stained with antibody cocktail of anti-CCR7 and anti-CXCR3 in PBS for 30 minutes at 37°C, then with an antibody cocktail of viability and surface markers in BD Brilliant Stain Buffer (BD Biosciences) for 30 minutes at room temperature. This was followed by a centrifugation wash in 2% FBS 2mM EDTA (in PBS), and fixation and permeabilisation of cells using CytoFix/CytoPerm (BD Biosciences) prior to intra-cellular staining with antibodies to functional markers for 30 minutes at room temperature. Cells were then washed in Perm/Wash (BD Biosciences) and fixed in 1% paraformaldehyde (Kimix) in PBS prior to acquisition on a LSRII flow cytometer (BD Biosciences; Supplementary Figure 1).

**Data Analysis Pipeline (PBMC-ICS cohort)**

***COMPASS Analysis***

We utilised flow cytometry to measure the expression of 5 functional markers, IFN-γ, TNF, IL-2, CD107 and CD154 by T cells in mycobacteria-stimulated PBMC. In addition to measuring expression of each functional marker, we also wanted to determine co-expression of every possible combination of these markers (n = 31). However, not all combinations would be expressed by M.tb-specific T cells nor present at frequencies higher than background. To determine which functional cell subsets were M.tb-specific, we utilised the COMPASS (version 1.20.1; https://github.com/RGLab/COMPASS) R package using R version 3.6.1 and RStudio Version 1.2.1335. COMPASS computes the posterior probability of each cell subset co-expressing functional markers based on cell counts in stimulated versus unstimulated conditions for each individual. COMPASS also computes two scores, the functionality (FS) and polyfunctionality (PFS) scores. The FS is defined as the proportion of antigen-specific subsets co-expressing functional markers detected among all possible ones. The PFS is similar, but it weighs the different subsets by their degree of functionality where subsets with higher degrees of functionality are favoured [2]. We conducted COMPASS analysis of CFP-10/ESAT-6, M.tb lysate and SEB-specific CD4 T cell subsets at all study visits. We only analysed subsets expressing different combinations of cytokines if they had a posterior probability threshold of ≥ 0.1 in at least 10 participants/visit for recent and remote infection. This arbitrary cut-off was used to ensure that subsets that were further analysed were all antigen-specific, higher than background and to minimise experimental artefacts. Functional subsets (booleans) that passed these criteria were then analysed after background subtraction using Pestle and SPICE. Regardless of posterior probability threshold, we calculated FS and PFS for each participant.

***CITRUS analysis***

We included 5 functional (IFN-γ, TNF, IL-2, CD154 and CD107) and 6 phenotypic (CD45RA, CCR7, CD27, KLRG-1, CXCR3 and HLA-DR) markers in our analyses, amounting to a total of 2048 potential co-expression combinations. The majority of these marker combinations may not be expressed by mycobacteria-specific CD4 T cells. To determine which functional and phenotypic combinations changed upon recent infection and which ones were different between recent and remote infection, we utilised CITRUS analysis to identify subsets that were significantly different between two groups or to define the minimum number of markers that can predict differences between two groups [3].

Definition of CITRUS clusters: We utilised the significance analysis of microarrays (SAM) association model to determine which co-expression patterns were significantly different between groups. We randomly down-sampled an equal number of events per participant and performed clustering characterisations based on marker abundance, which identifies differences based on the proportional contribution of each marker to the defined cluster. CITRUS clustering was performed based on memory markers, CD45RA, CCR7, CD27 and KLRG1, and Th1 cytokines IFN-γ, TNF and IL-2. CD107, CD154, CXCR3 and HLA-DR were not used to define the final CITRUS clusters because preliminary analysis showed that inclusion of these markers was not useful to further delineate the clusters in addition to the other 7 markers. CITRUS outputs data in the form of CITRUS trees, coloured by parameter abundance, highlights differentially expressed clusters that passed a FDR threshold of < 0.05 (Supplementary Figure 3 A-B) and depicts composition of clusters using histograms (Supplementary Figure 3 C).

Confirmation of CITRUS clusters in FlowJo: To confirm the memory and functional marker composition of clusters identified to be differentially abundant, we exported each CITRUS cluster that passed the FDR threshold of 0.05 and concatenated the FCS files belonging to the same cluster from all participants in FlowJo. Clusters were then manually reanalysed to determine the minimum set of markers whose combined expression "reproduced" each CITRUS cluster (Supplementary Figure 4). Upon confirmation of cluster composition, we manually gated cell subsets representative of these clusters on individual responder FCS files and used this data to present the results described.

**CITRUS Analysis results**

CITRUS analyses identified 13 M.tb lysate-specific CD4 T cell clusters that were expressed at significantly different abundance levels upon recent M.tb infection, and 4 M.tb lysate-specific and 3 CFP-10/ESAT-6-specific CD4 T cell clusters that were significantly different between recent and remote infection (FDR < 0.05; Supplementary Table 5). Before confirming cluster identity by manual gating, we compared histogram expression patterns of differentially expressed clusters to ensure that there were no redundant populations. This quality control step reduced the number of distinct differentially expressed populations that were confirmed using manual gating in Flow Jo (Supplementary Table 6; Figure 3). Over 90% of cells in Cluster 2570 were CD45RA+ memory cells, however proportions of CD45RA+ memory cells constitute <10% of total memory cells. We therefore separated cluster 2570 into CD45RA+ and CD45RA- cell subsets. We noted that clusters 2566 and 2559; and 2238 and 2219 had similar expression patterns and could be defined as the same populations (Supplementary Table 6; Figure 3).

### Figures

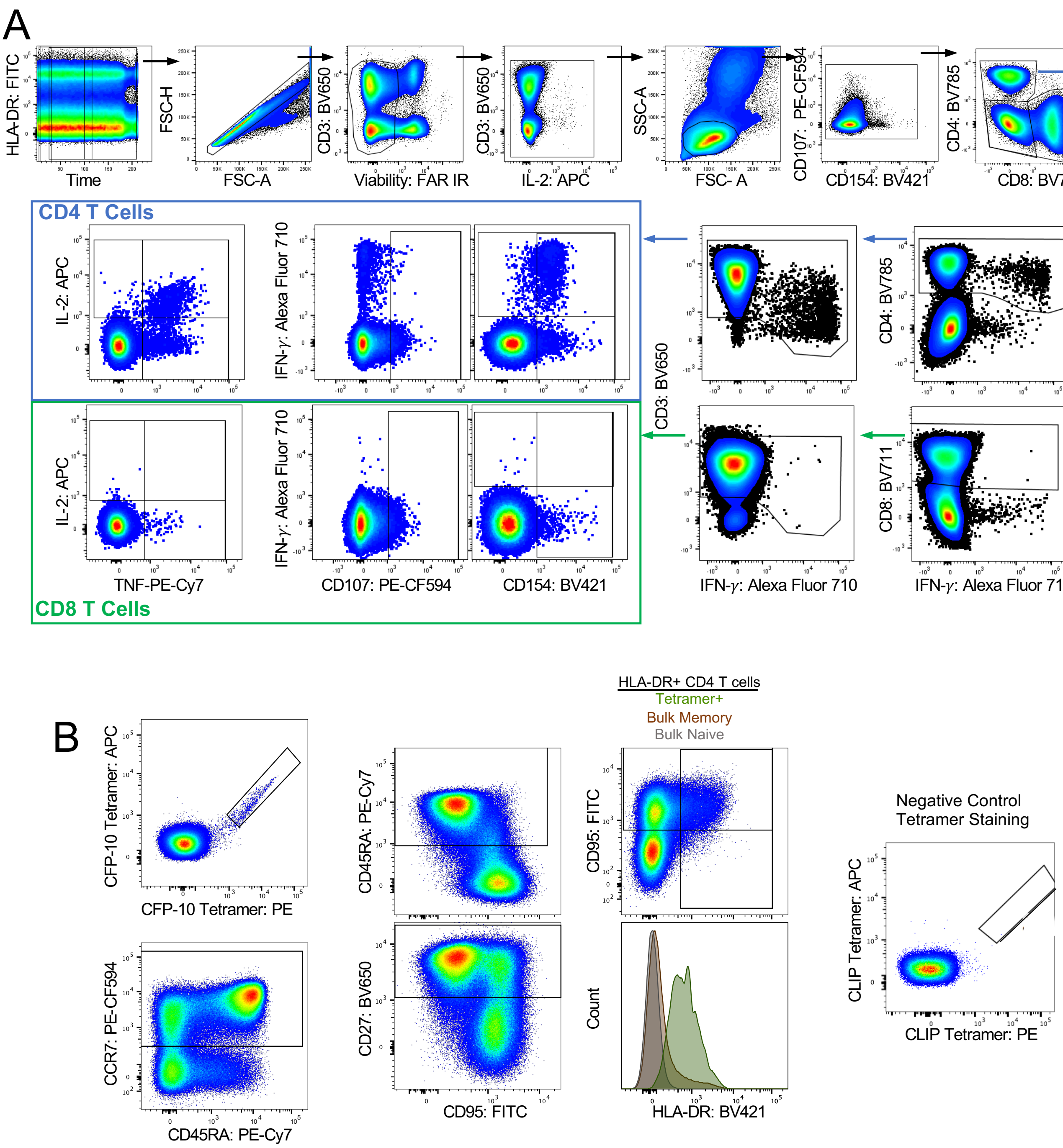


**Supplementary Figure 1: Gating strategy used to identify M.tb-specific CD4+ T cells. (A)** To identify M.tb-responsive T cells in the PBMC-ICS assay, we first gated on total cells based on fluorescence consistency over acquisition time (time gate), followed by exclusion of cell doublets, dead cells and antibody aggregates. We then gated on total lymphocytes and excluded additional staining artefacts. We noted major down-regulation of CD3 expression on cytokine-expressing functional cells. Thus, to ensure that we correctly classified all cytokine+ T cells as CD4+ or CD8+ CD3+ T cell, we first gated on CD4+ (after excluding CD8+ cells) and CD8+ (after excluding CD4+ cells) T cells and then gated on CD3+ T cells. Functional markers (IFN-γ, TNF, IL-2, CD154 and CD107) were then gated independently on CD4+CD3+ (blue box) and CD8+CD3+ (green box) T cells. (B) We co-stained samples with HLA class II tetramer reagents that were conjugated to 2 different fluorophores )APC and

-PE) to identify M.tb-tetramer+ CD4 T cells with high specificity. Gates for activation (HLA-DR) and memory markers (CD45RA, CCR7, CD27 and CD95) were set on total CD4+ T cells. Bulk naïve cells were defined as CD45RA+CCR7+CD27+CD95- CD4 T cells, while bulk memory cells were defined as non-naïve CD4 T cells. The gate identifying HLA-DR+ cells was set on total CD4 T cells and then applied to M.tb-tetramer+, bulk naïve and bulk memory T cells. Full gating strategy was previously published (Mpande *et al.,* 2018).

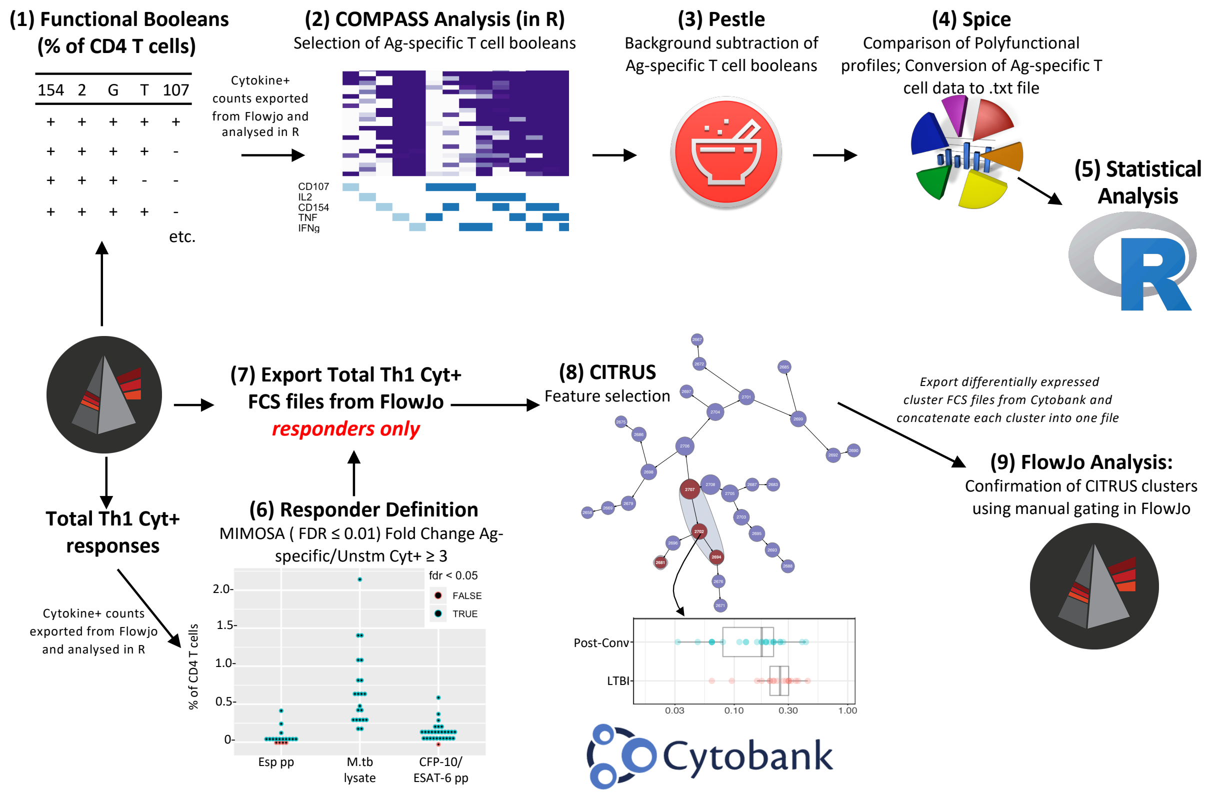


**Supplementary Figure 2: PBMC-ICS Analysis Pipeline**. (1) All combinations of functional subsets (booleans of IFN-γ±TNF±IL-2±CD154±CD107±; n=31) were defined using Flowjo and (2) antigen-specific CD4 T cell subsets were identified using COMPASS in R. (3) To account for non-specific responses, we subtracted background (unstimulated) frequencies for each Boolean subset in Pestle. (4) The distribution of polyfunctional cell subsets (pie-charts) across groups were compared in SPICE. (5) Statistical comparisons of functional booleans were conducted in R. In parallel, (6) responders to antigenic stimulation were defined on total cytokine+ CD4 T cells (expressing any combination of IFN-γ, TNF or IL-2) that passed a MIMOSA FDR threshold of ≤ 0.01 and had a fold change of antigen-specific signal over unstimulated (≥ 3). (7) Total Th1 cytokine+ FCS files for responders only were exported from FlowJo and (8) imported into Cytobank to determine differentially expressed clusters between two groups using CITRUS. (9) Identity of clusters defined in CITRUS was confirmed using manual gating in FlowJo and the final gating strategy for each cluster was applied to individual responders.

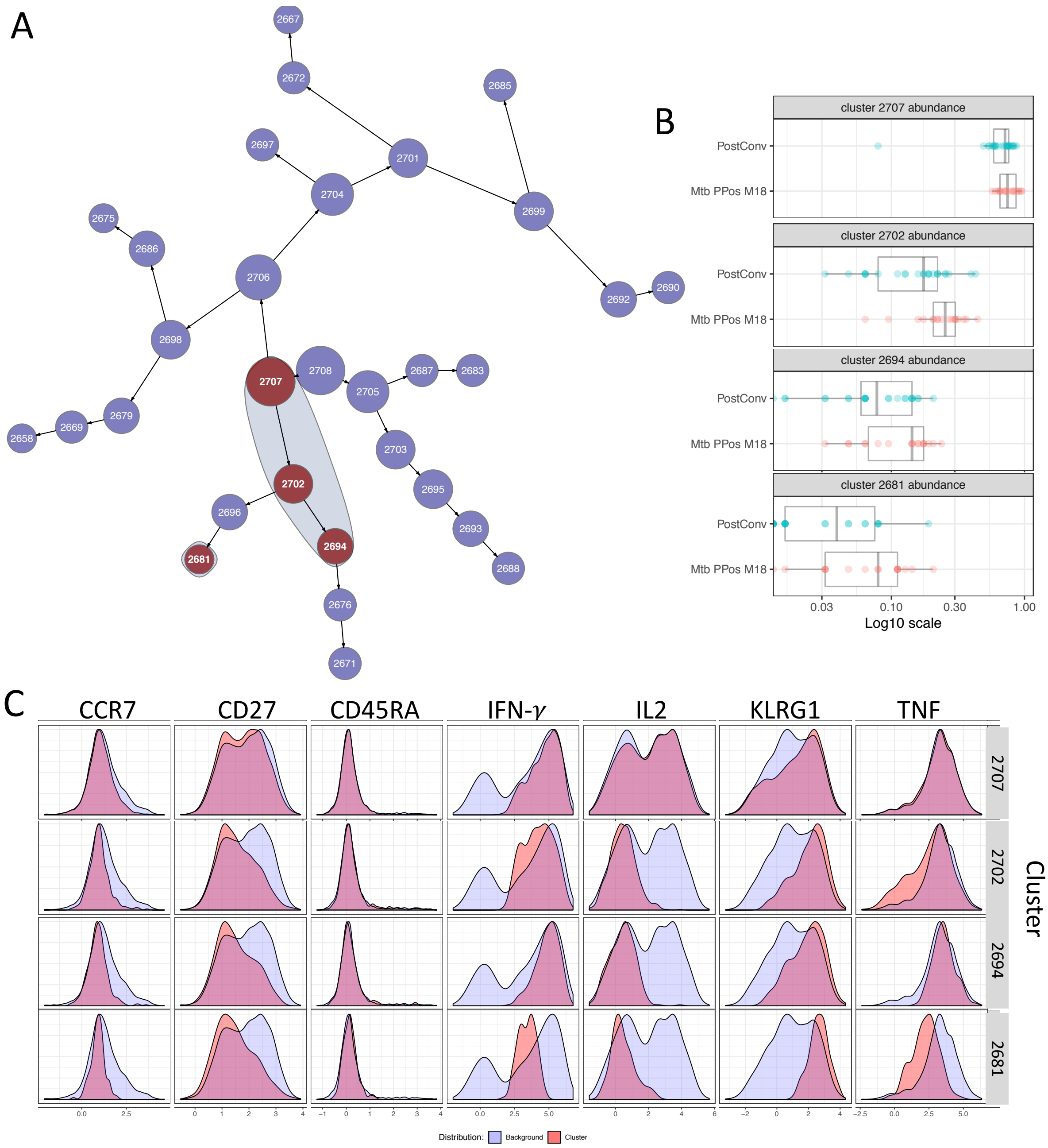


**Supplementary Figure 4: Recent vs remote TBI CITRUS analysis result.** Representative example of CITRUS-defined clustering represented as a (A) CITRUS tree. Shaded clusters represent clusters that are differentially expressed with an FDR threshold of < 0.05 between recent and remote infection. Differentially expressed clusters are also represented in (B) graphical format depicting expression by each participant. (C) Expression of each memory/functional marker used to define the cluster (salmon) compared to total Th1 cells (light purple) are depicted in overlaid histograms.

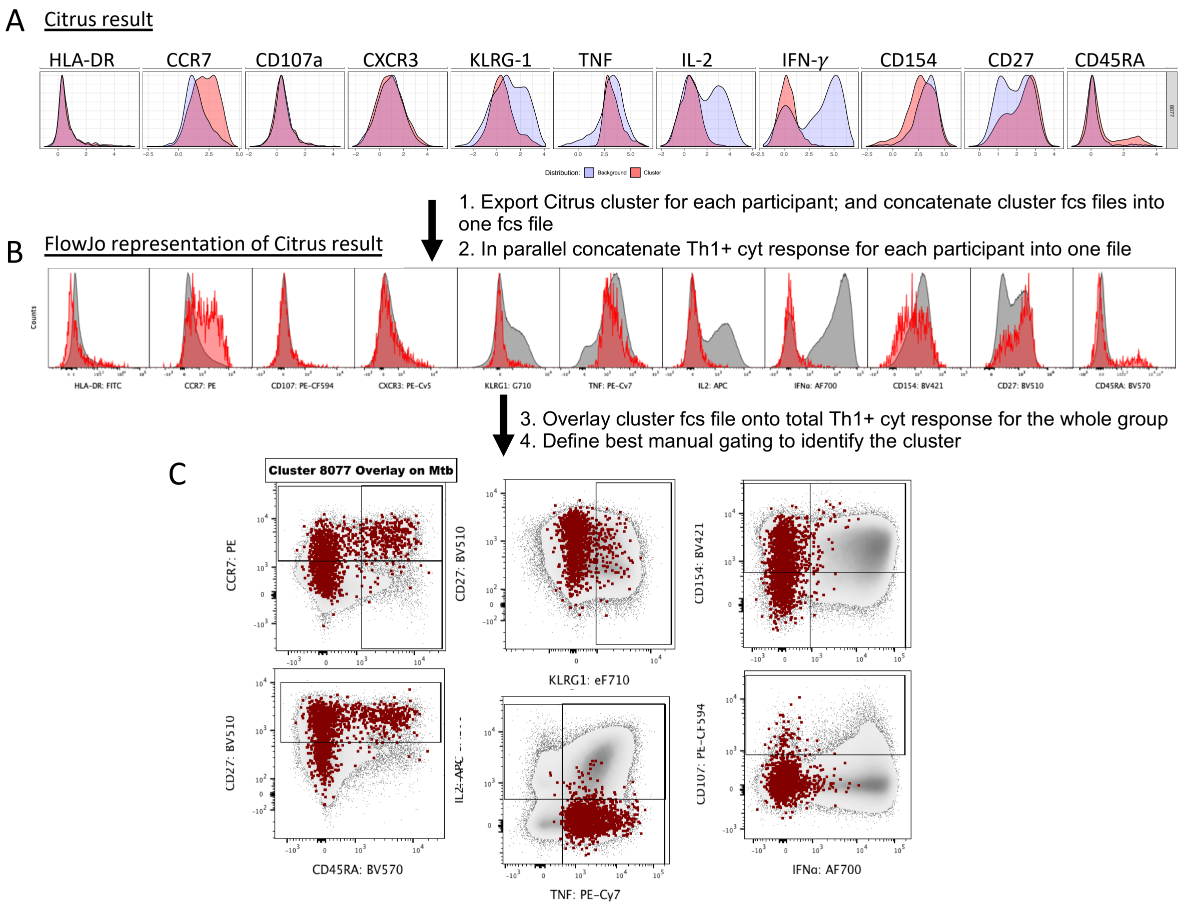


**Supplementary Figure 4: Analysis pipeline used to confirm CITRUS cluster co-expression profiles.** (A) CITRUS clusters were exported from Cytobank and concatenated into one file using Flowjo. In parallel, total Th1+ cytokine (cyt) responses for each participant were also concatenated into one file (representing Th1+ cyt response for all participants). (B) Concatenated citrus cluster (red) was overlaid on total Th1+ cyt response (grey) to validate CITRUS modelling of marker expression. (C) Concatenated cluster events were overlaid on total Th1+ cyt, visualised by conventional bi-variate plots, and expression levels of memory markers and cytokines were used to define the best gating strategy to manually validate the clusters based on co-expression of CD45RA, CCR7, CD27, KLRG1, IFN-γ, TNF and IL-2.

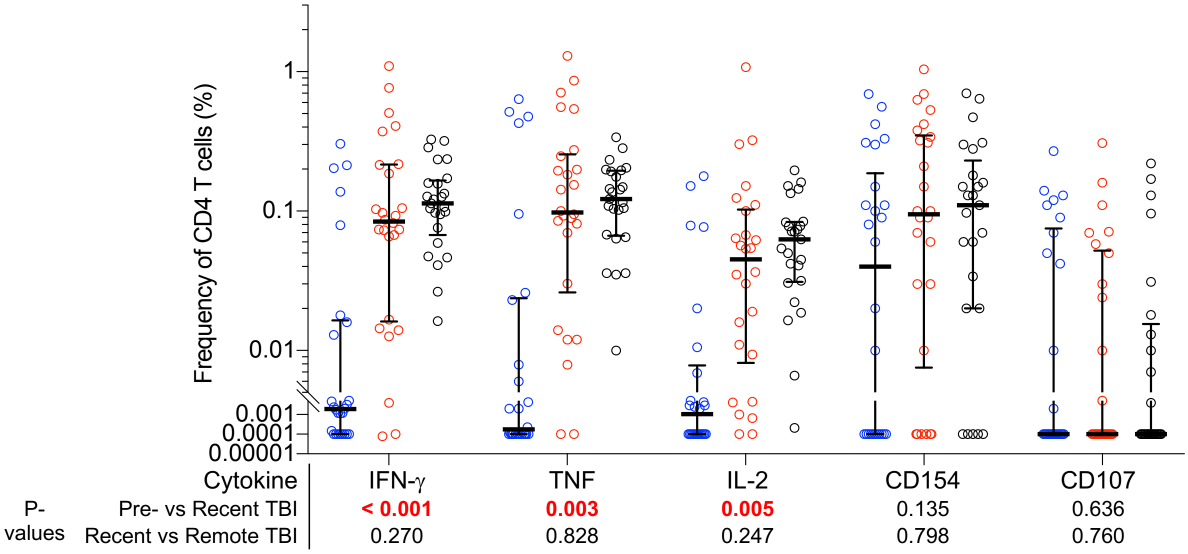


**Supplementary Figure 5: Acquisition of infection is associated with induction of M.tb-specific functional CD4 T cell responses.** Frequencies of background subtracted CFP-10/ESAT-6-specific IFN-γ+ TNF+, IL-2+, CD154+ and CD107+ CD4 T cells detected before (pre-TBI, blue, n=26), after (recent TBI, red, n=26) and during remote TBI (black symbols, n=25). P-values were calculated using the Wilcoxon-signed rank for paired (pre-TBI *versus* recent TBI) or the Mann-Whitney U test for unpaired (recent *versus* remote TBI) comparisons and corrected for multiple comparisons using the Benjamini–Hochberg method with a false discovery rate (FDR) of 0.05. P-values highlighted in **red, bold text** were considered significant. Values less than 0.0001 were set to 0.0001 to allow visualisation on a logarithmic scale.

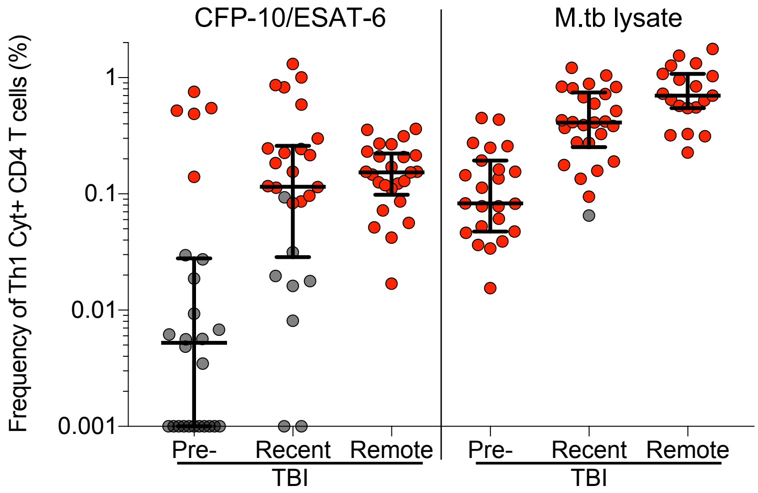


**Supplementary Figure 6: TBI is associated with a M.tb-specific high responder rate.** Frequencies of background subtracted CFP-10/ESAT-6-specific (n: pre-TBI =26, recent TBI = 26, remote TBI = 25) and M.tb lysate-specific (n: pre-TBI = 23, recent TBI= 26, remote TBI=19) total Th1 (IFN-γ±TNF±IL-2±) cytokine+ CD4 T cells detected in the biomarker discovery cohort. Symbols depict responder (red) and non-responder (grey) participants.

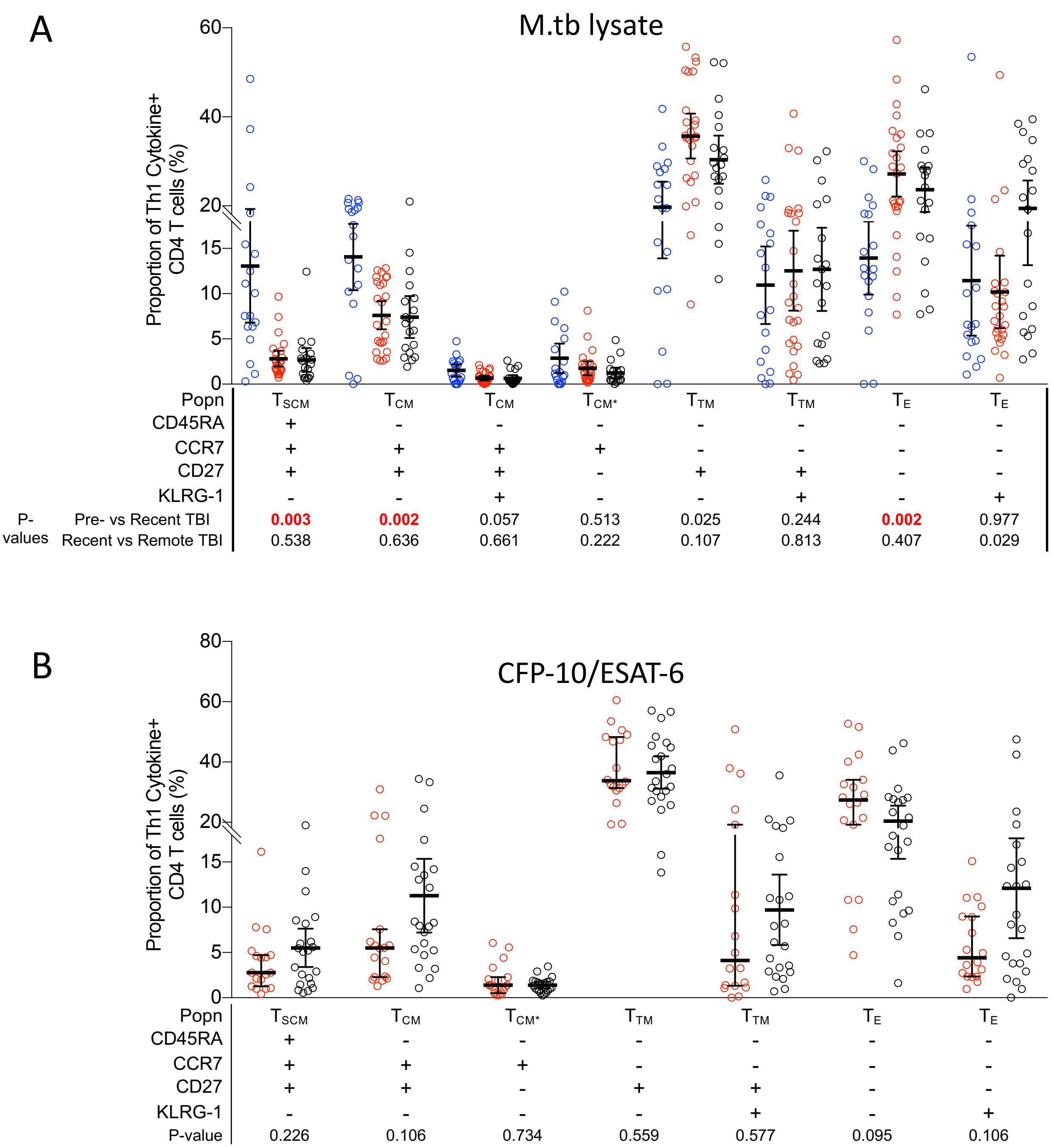


**Supplementary Figure 7: Recent TBI is associated with a decrease in less differentiated T cell subsets, and predominantly differentiated memory T cell subsets, which are similar between recent and remote TBI.** Proportions of (A) M.tb lysate-specific and (B) CFP-10-ESAT6-specific memory profiles detected before (Pre-TBI, blue, M.tb lysate, n=16), after (recent TBI, red, CFP-10/ESAT-6, n=18; M.tb lysate, n=26), and during remote TBI (black, CFP-10/ESAT-6, n=22; M.tb lysate, n=19). Statistical analysis was conducted as in Supplementary Figure 5. P-values highlighted in **red, bold text** were considered significant.

### Tables

#### Supplementary Table 1: Demographic characteristics of participants in each sub-group.

| | Tetramer Cohort | Biomarker Discovery cohort | |
|---|---|---|---|
| | | **Remote TBI** | **Recent TBI** |
| **n** | 11 | 30 | 30 |
| **Female gender (%)** | 7 (64%) | 21 (70%) | 19 (63%) |
| **Median age in years (IQR)** | 17 (15.5-17) | 15 (14-16) | 15 (14-16) |
| **Ethnicity: Coloured** | 11 | 26 | 29 |
| **Ethnicity: Black** | 0 | 4 | 1 |

*Participant samples selected for the tetramer cohort were based on their HLA types that matched available MHC II tetramers and are the same participants utilised to generate some of the results described in Mpande *et al.,* 2018

#### Supplementary Table 2: PBMC-ICS Flow Cytometry Panel

| Marker | Role | Fluorochrome | Clone | Manufacturer | Cat. Number (RRID) |
|---|---|---|---|---|---|
| CD3 | Lineage | BV650 | UCHT1 | BD Biosciences | 563852 (AB_2744391) |
| CD4 | Lineage | BV785 | OKT4 | BioLegend | 317442 (AB_2563242) |
| CD8 | Lineage | BV711 | RPA-T8 | BioLegend | 301044 (AB_2562906) |
| CCR7 | T cell differentiation | PE | 150503 | BD Biosciences | 560765 (AB_2033949) |
| CD27 | T cell differentiation | BV510 | L128 | BD Biosciences | 563092 (AB_2313577) |
| CD45RA | T cell differentiation | BV570 | HI100 | eBioscience | 304132 (AB_2563813) |
| KLRG-1 | T cell differentiation | PercP-eFlour710 | 13F12F2 | eBioscience | 46948842 (AB_2573889) |
| CXCR3 | $T_{SCM}$; Homing | PE-Cy5 | 1C6/CXCR3 | BD Biosciences | 551128 (AB_394061) |
| HLA-DR | Activation | FITC | L243 | BD Biosciences | 307604 (AB_314682) |
| CD107 | Function | PE-CF594 | H4A3 | BD Biosciences | 562628 (AB_2737686) |
| CD154 (CD40L) | Function | BV421 | TRAP-1 | BD Biosciences | 563886 (AB_2738466) |
| IFN-γ | Function | Alexa Fluor 700 | B27 | BD Biosciences | 557995 (AB_396977) |
| IL-2 | Function | APC | MQ1-17H12 | BD Biosciences | 554567 (AB_398571) |
| TNF | Function | PE-Cy7 | Mab11 | BioLegend | 25734982 (AB_469686) |
| Live/Dead | Viability | Near IR (APC-H7) | N/A | Life Technologies | L34976 |

**Supplementary Table 3: HLA-class II tetramer-peptide cognates**

| Tetramer | HLA Sequence | Protein | Peptide Sequence | Participants (n) |
|---|---|---|---|---|
| M.tb | DRB1*04:01 | CFP-10$_{71-85}$ | EISTNIRQAGVQYSR | 5 |
| M.tb | DRB5*01:01 | CFP-10$_{51-65}$ | AQAAVVRFQEAANKQ | 4 |
| M.tb | DQB1*06:02 | ESAT-6$_{31-45}$ | EGKQSLTKLAAAWGG | 3 |
| CLIP | DRB1*04:01; DRB5*01:01; DQB1*06:02 | CLIP$_{87-101}$ | PVSKMRMATPLLMQA | 12* |

Dual tetramer staining with identical tetramers conjugated to -APC and -PE were performed for all samples.
*Each M.tb-specific tetramer staining had a corresponding control (CLIP)-tetramer staining that was performed in parallel.

**Supplementary Table 4: Tetramer Cohort Flow Cytometry Panel**

| Marker | Role | Fluorochrome | Clone | Manufacturer | Cat. Number (RRID) |
|---|---|---|---|---|---|
| CD3 | Lineage | Alexa Fluor 700 | UCHT1 | BD Biosciences | 557943 (AB_396952) |
| CD4 | Lineage | BV785 | L200 | BioLegend | 317442 (AB_2563242) |
| CD8 | Exclusion | BV510 | RPA-T8 | BioLegend | 301048 (AB_2561942) |
| CD14 | Exclusion | BV510 | M5E2 | BioLegend | 301842 (AB_2561946) |
| CD19 | Exclusion | BV510 | SJ25C1 | BD Biosciences | 562947 (AB_2737912) |
| CCR7 | T cell differentiation | PE-CF594 | 150503 | BD Biosciences | 562381 (AB_11153301) |
| CD27 | T cell differentiation | BV650 | L128 | BD Biosciences | 563228 (AB_2744352) |
| CD45RA | T cell differentiation | PE-Cy7 | HI100 | eBioscience | 25045842 (AB_1548774) |
| CD95 | T cell differentiation | FITC | DX2 | BioLegend | 305606 (AB_314544) |
| HLA-DR | Activation | BV421 | L243 | BioLegend | 307636 (AB_2561831) |
| Live/Dead | Viability | Aqua (BV510) | N/A | Thermo-Fisher Scientific | L34957 |

**Supplementary Table 5 Number of differentially (FDR < 0.05) expressed clusters identified between 2 groups using CITRUS.**

| Ag Specificity | TBI Comparison | Total Clusters | Differentially expressed (DE) clusters | DE distinct populations | Cell populations confirmed by Manual Gating in FlowJo |
|---|---|---|---|---|---|
| M.tb lysate | Pre vs Recent | 29 | 13 | 6 | 6 |
| | Recent vs Remote | 32 | 4 | 2 | 2 |
| CFP-10/ESAT-6 | | 32 | 3 | 3* | 2 |

*3 differentially expressed distinct populations were identified following CITRUS analysis. Investigation of memory and functional marker composition revealed that two of these clusters (which branched off different nodes) could be classified as one cluster as they had similar expression patterns. We therefore only confirmed manually confirmed 2 distinct populations.

**Supplementary Table 6**

| Antigen-specificity | TBI Comparison | Cluster | CD45RA | CCR7 | CD27 | KLRG-1 | IFN-γ | TNF | IL-2 |
|---|---|---|---|---|---|---|---|---|---|
| M.tb lysate | Pre- vs Recent | 2565 | 40.6 | **82.7** | **81.2** | **8.27** | **8.27** | **14.3** | **99.2** |
| | | 2570 | **94.8** | **99.3** | **97.4** | **10.5** | **28.8** | **85.6** | **17** |
| | | 2566 | **0.6** | **15** | 55.7 | **3.59** | **90.4** | **98.2** | **93.4** |
| | | 2559 | **0** | **2.79** | 49.2 | **0** | **100** | **96.6** | **82.7** |
| | | 2567 | **0** | **4.78** | 53 | **3.04** | **95.7** | **91.3** | **3.04** |
| | | 2551 | **0** | **3.33** | **73.8** | **74.8** | **100** | **100** | **98.1** |
| | Recent vs Remote | 2694 | **3.83** | **2.09** | 36.6 | 50.2 | **100** | **99.3** | **1.05** |
| | | 2681 | **0** | **0** | **33.1** | **84.5** | **99.3** | **66.9** | **5.41** |
| CFP-10 /ESAT-6 | | 2219# | **0** | **9.46** | 39.2 | **0** | **0** | **100** | **9.46** |
| | | 2238# | **0.51** | **13.1** | 51 | **8.08** | **8.08** | **99.5** | **4.55** |
| | | 2243 | **0.28** | **33.8** | **70.2** | **4.55** | **4.55** | **99.7** | **3.12** |

Numbers denote expression of each marker as percentage (%) of total M.tb-specific CD4 T cells in each cluster. Markers expressed in less than 35% and more than 65% of total M.tb-specific cells are denoted in blue and red, respectively, and were used to define the cluster using manual gating. These arbitrary cut-offs were chosen because it enabled manual gating of clusters that were most similar to the histogram depicting cluster composition. #: clusters 2219 and 2238 had similar expression patterns but branched off 2 different CITRUS tree nodes, as a result only 2 distinct populations were confirmed for recent vs remote TBI (CFP-10/ESAT-6 specific CD4 T cell) comparison: 2219/38 and 2243.

**Supplementary Table 7**

| CD4 T cell subset 1 | | | CD4 T cell subset 2 | | | Roc test p-value |
|---|---|---|---|---|---|---|
| **Subset** | ***Stim*** | ***AUCROC (95% CI)*** | **Subset** | ***Stim*** | ***AUCROC (95% CI)*** | |
| HLA-DR% of IFN-γ+ | P10ES6 | 0.94 (0.83-1.00) | HLA-DR % of IFN-γ+ | M.tbL | 0.83 (0.69-0.96) | 0.198 |
| | | | HLA-DR % of IFN-γ+TNF+ | P10ES6 | 0.94 (0.84-1.00) | 0.332 |
| | | | HLA-DR % of IFN-γ+TNF+ | M.tbL | 0.82 (0.68-096) | 0.197 |
| | | | HLA-DR % of Th1 Cyt+ | P10ES6 | 0.92 (0.81-1.00) | 0.227 |
| | | | HLA-DR % of Th1 Cyt+ | M.tb | 0.83 (0.70-0.96) | 0.201 |
| | M.tbL | 0.83 (0.69-0.96) | HLA-DR % of IFN-γ+TNF+ | P10ES6 | 0.94 (0.84-1.00) | 0.180 |
| | | | HLA-DR % of IFN-γ+TNF+ | M.tbL | 0.82 (0.68-096) | 1.000 |
| | | | HLA-DR % of Th1 Cyt+ | P10ES6 | 0.92 (0.81-1.00) | 0.295 |
| | | | HLA-DR % of Th1 Cyt+ | M.tbL | 0.83 (0.70-0.96) | 0.762 |
| | | | IFN-γ+TNF+IL-2- $T_E$ | M.tbL | 0.77 (0.62-0.92) | 0.504 |
| | | | IFN-γ+TNF+IL-2- KLRG- 1+ $T_E$ | M.tbL | 0.74 (0.58-0.90) | 0.333 |
| | | | FDS | M.tbL | 0.72 (0.56-0.88) | 0.272 |
| HLA-DR % of IFN-γ+TNF+ | P10ES6 | 0.94 (0.84-1.00) | HLA-DR % of IFN-γ+TNF+ | M.tbL | 0.82 (0.68-096) | 0.197 |
| | | | HLA-DR % of Th1 Cyt+ | P10ES6 | 0.92 (0.81-1.00) | 0.188 |
| | | | HLA-DR % of Th1 Cyt+ | M.tbL | 0.83 (0.70-0.96) | 0.182 |
| | M.tbL | 0.82 (0.68-096) | HLA-DR % of Th1 Cyt+ | P10ES6 | 0.92 (0.81-1.00) | 0.294 |
| | | | HLA-DR % of Th1 Cyt+ | M.tbL | 0.83 (0.70-0.96) | 0.767 |
| | | | IFN-γ+TNF+IL-2- $T_E$ | M.tbL | 0.77 (0.62-0.92) | 0.481 |
| | | | IFN-γ+TNF+IL-2- KLRG- 1+ $T_E$ | M.tbL | 0.74 (0.58-0.90) | 0.339 |
| | | | FDS | M.tbL | 0.72 (0.56-0.88) | 0.283 |
| HLA-DR % of Th1 Cyt+ | P10ES6 | 0.92 (0.81-1.00) | HLA-DR % of Th1+ | M.tbL | 0.83 (0.70-0.96) | 0.302 |
| | | | IFN-γ+TNF+IL-2- $T_E$ | M.tbL | 0.77 (0.62-0.92) | 0.104 |
| HLA-DR % of Th1 Cyt+ | M.tbL | 0.83 (0.70-0.96) | IFN-γ+TNF+IL-2- $T_E$ | M.tbL | 0.77 (0.62-0.92) | 0.457 |
| | | | IFN-γ+TNF+IL-2- KLRG- 1+ $T_E$ | M.tbL | 0.74 (0.58-0.90) | 0.333 |
| | | | FDS | M.tbL | 0.72 (0.56-0.88) | 0.252 |
| IFN-γ+ TNF+IL-2- $T_E$ | | 0.77 (0.62-0.92) | IFN-γ+TNF+IL-2- KLRG- 1+ $T_E$ | M.tbL | 0.74 (0.58-0.90) | 0.636 |
| | | 0.77 (0.62-0.92) | FDS | M.tbL | 0.72 (0.56-0.88) | 0.540 |
| IFN-γ+TNF+IL-2- KLRG- 1+ $T_E$ | | 0.74 (0.58-0.90) | FDS | M.tbL | 0.72 (0.56-0.88) | 0.818 |

Abbreviations: Stimulation: Stim; CFP-10/ESAT-6: P10/ES6; M.tb lysate: M.tbL.
